# Open-Source Light Calibration System for Hyperbilirubinemia Phototherapy Treatments

**DOI:** 10.1101/2025.08.01.25332669

**Authors:** Joshua T.M. Givans, Augustine Waswa, June Madete, Joshua M. Pearce

## Abstract

Neonatal jaundice or neonatal hyperbilirubinemia is a common medical condition impacting newborns and pathological jaundice if left untreated leads to neurological encephalopathy and/or death. The majority of pathological jaundice cases occur in low and middle- income countries (LMIC). Phototherapy has been determined to be the safest and most effective treatment for jaundice. Although inexpensive light emitting diodes are available on the market, commercial phototherapy devices are expensive (∼US$2,000), which creates a barrier to access for these devices in LMIC. Efforts to construct cost effective phototherapy units have been implemented in the past but need a method to validate the intensity and wavelength of light received by the infant at a distance away from the source. To enable low-cost phototherapy units to be used clinically, this study provides an open source, low-cost, distributed manufacturing approach to create a light sensor to calibrate phototherapy units. This instrument is a necessary component of any open source phototherapy treatment used in a clinical setting. This novel instrument was validated by comparing its irradiance and wavelength reading to the commercially calibrated Ocean Insight UV-VIS spectrometer under varying lighting conditions including that of the existing Datex-Ohmeda Giraffe Spot PT Lite phototherapy equipment accessible through Victoria Children’s Hospital Neonatal Care Ward in London, Ontario and Kiambu County Hospital in Kenya. The results of this study have demonstrated that for under US$150, a phototherapy calibration device can be constructed capable of measuring up to 200uW/cm^2^/nm with an accuracy of 98.6% and detect the peak wavelength within +/- 12.5 nm. It can be concluded that 3D printed open- source irradiance meters are a viable option for calibrating phototherapy units in LMIC to treat hyperbilirubinemia.

## Introduction

Neonatal jaundice or neonatal hyperbilirubinemia is one of the most common medical conditions to affect newborns (1). Roughly 60% of all newborn infants are diagnosed with jaundice (2,3). Most of these cases are of a milder condition- physiological jaundice- and resolves on its own (2,3).

Pathological jaundice, however, is a far more serious condition, which if left untreated leads to neurological encephalopathy and/or death (2,3). Pathological jaundice accounts for ∼25% of diagnoses, which still represents an enormous number of impacted newborns (2,3). The majority of cases have been observed to occur in low and middle- income countries (LMIC) (2,4,5). The increased cases of jaundice in LMIC can be attributed to a lack of appropriate medical technology and sustainable processes to treat the condition (5).

The development of neonatal jaundice occurs as a result of the infant’s inability to breakdown and process hemoglobin in the blood (3). Bilirubin, a compound derived from hemoglobin, can be difficult for an infant to excrete for a variety of reasons including: increased production of bilirubin, deficient conjugation of bilirubin, impaired hepatic uptake, and/or enhanced enterohepatic circulation of bilirubin (3). Phototherapy, exchange blood transfusions and intravenous immunoglobulin have been identified as effective treatments for overcoming an infant’s inability to breakdown and process hemoglobin in the blood that causes jaundice (3,6,7). Phototherapy is the most common method to treat hyperbilirubinemia while exchange transfusion is reserved for the most severe cases (8).

Phototherapy has been determined to be the safest and most effective treatment for jaundice (1,3,6,9–17). Light of the appropriate wavelength and intensity isomerises bilirubin in the blood stream into compounds more readily excreted by the body (16–18). The recommended characteristics of an effective phototherapy device are: peak wavelength between 460 – 490 nm, irradiance > 30uW/cm^2^/nm and effective treatment area > 2000cm^2^ (9,14,19).

The quality of treatment varies with the intensity of light received and as a case study in Nigerian hospitals presents: the mean irradiance received by the infant was below optimal levels (10). This is not an isolated event, it is known that the intensity of phototherapy devices decay over time (20). Simple and affordable methods to maintain the standard of treatment in LMIC are in need (10,20). A typical irradiance meter designed to maintain the effectiveness of a phototherapy unit typically costs upwards of $2,000 (Table 1). The high cost creates a barrier to access to these devices in LMIC. The United Nations Children’s Fund (UNICEF) has developed a target product profile which outlines the requirement for a phototherapy unit which is both an effective treatment for jaundice and a cost effective solution in LMIC (4,19). Included in this outline is the requirement that an irradiance meter be made available alongside the unit such that a clinician can verify the appropriate dosage is administered to the infants.

**Table 1.** Market Survey of Irradiance Meters.

| Device Name | Cost (USD) | Source |
| --- | --- | --- |
| UNICEF Phototherapy Irradiance Meter | \$ 1,679.00 | (21) |
| DALE40 Phototherapy Radiometer | \$ 3,201.38 | (22) |
| ILT750 Bili Light Meter | \$ 1,999.00 | (23) |

Efforts to construct cost effective phototherapy units have been implemented in the past. A solar powered phototherapy device (24) used an LDR photo resistor to determine the intensity of light emitted by the source. This method may accurately determine the intensity of light at the source but cannot provide information about the intensity of light received by the infant at a distance away from the source. Furthermore, this method is unable to verify that the wavelength of the source falls within the recommended range. The open source irradiance meter described in this paper intends to build upon this work by implementing a separate device to validate the intensity and wavelength at the location of the infant.

One approach to reducing costs for scientific (25–27) and medical equipment (28–30) is the use of open hardware (31). In the open hardware approach digital designs of parametric hardware are shared freely on the web where anyone can download and replicate the device (32). As the cost of replication is made up of only materials and processing costs, open hardware can generally be manufactured for one tenth the cost of proprietary hardware (33). Open source hardware thus provides the opportunity for access to needed treatments in LMICs and other low-resource settings (34) if an open hardware design is published. In the medical context open hardware must also be calibrated for clinical acceptance (35). No such open design or calibration standards have been provided in the literature.

To provide for this need, this paper provides an open source, distributed manufacturing approach to create a light sensor to calibrate phototherapy units. This instrument is a necessary component of any open source phototherapy treatment used in a clinical setting. This instrument aims to validate both the wavelength of light produced and the intensity of light received by the patient. It is necessary that this instrument is both low cost and appropriately calibrated if it is to be used alongside phototherapy devices in LMIC. The instrument was validated by comparing its irradiance and wavelength reading to the commercially calibrated Ocean Insight UV-VIS spectrometer (36) under varying lighting conditions including that of the existing Datex-Ohmeda Giraffe Spot PT Lite phototherapy equipment (37) accessible through Victoria Children’s Hospital neonatal care ward in London, Ontario and Kiambu county hospital in Kenya.

## Materials and Methods

This study details an approach to calibrate an open source irradiance meter which in turn can be used to calibrate phototherapy units. The aim is to achieve accurate measurements of the light intensity, +/- 0.01 uW/cm^2^ and within a range of 0.01 – 150 uW/cm^2^ as described by the UNICEF design catalogue product requirements (21). Two designs were pursued: 1) using the APDS-9960 sensor (38) built into the Arduino BLE sense (39) shown in Figure 1 and 2) using the AS7265x sensor (40) shown in Figure 2.

**Figure 1.**
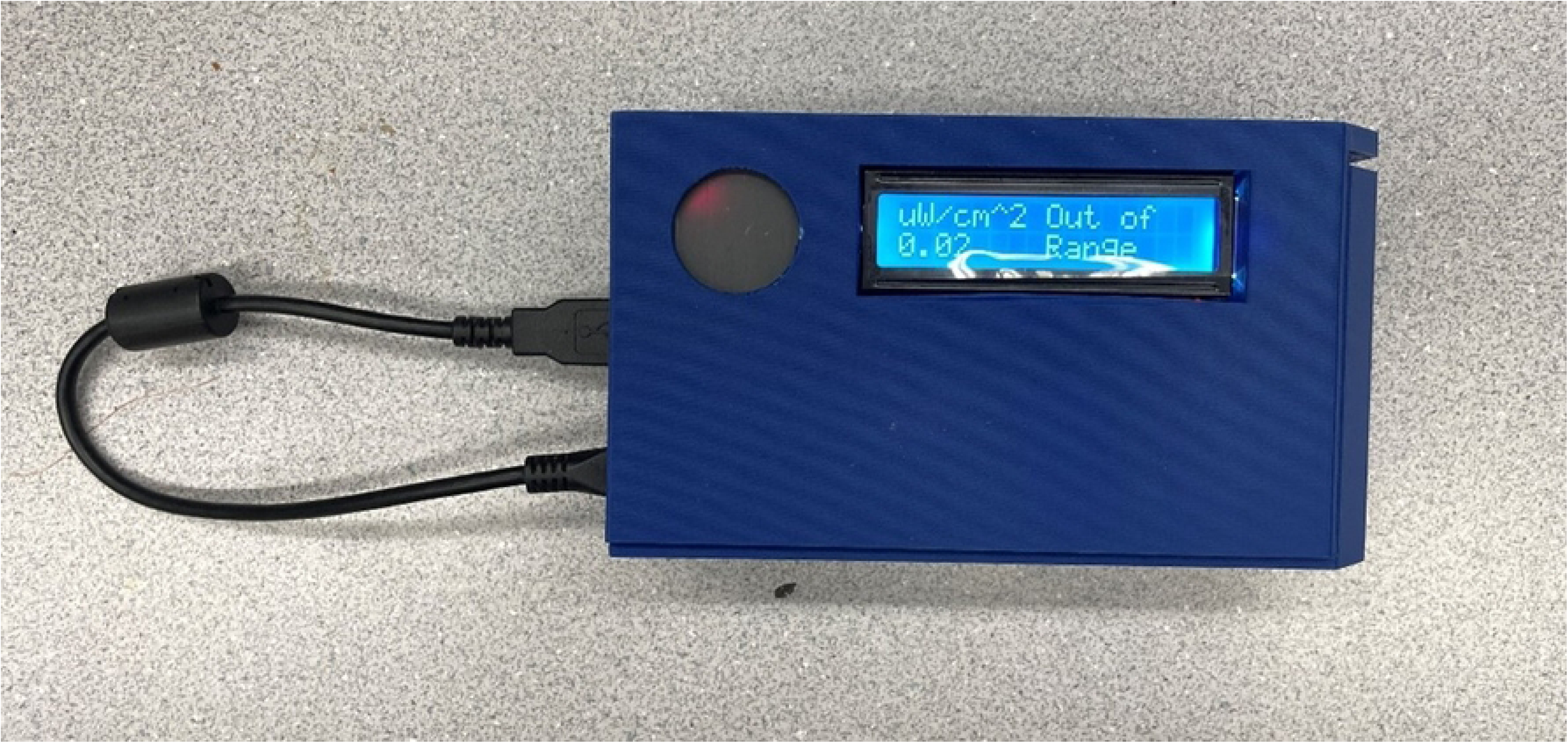
Built-in ADPS-9960 Sensor

**Figure 2.**
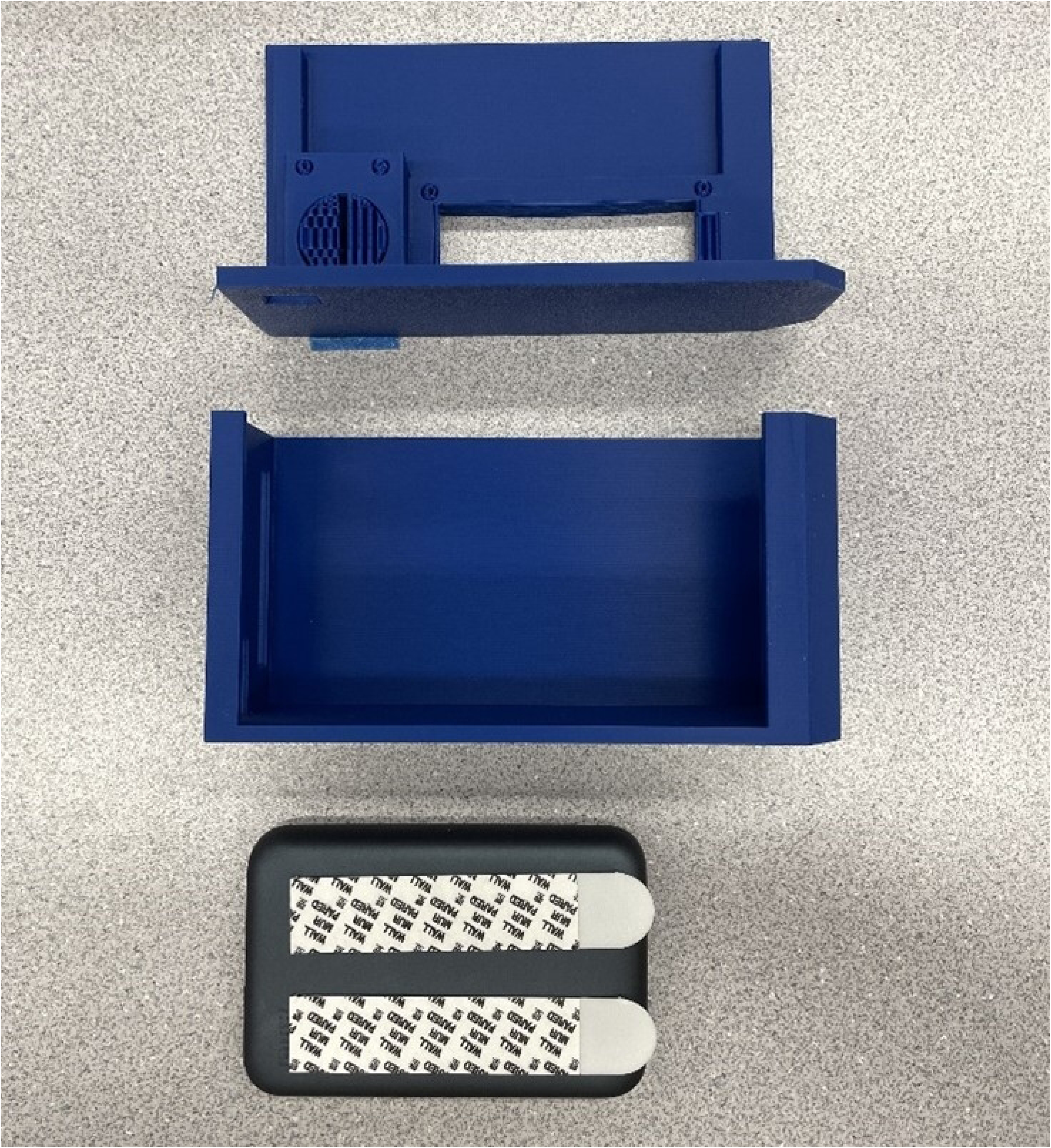
AS7256x External Sensor

Build instructions to create one independently can be found in Appendix A1 and A2, respectively. Both devices were exposed to an array of LED’s emitting at peak wavelength of 455 nm to simulate clinical settings. A calibrated spectrometer was used as the reference to which the irradiance meter was calibrated.

### Calibrate Irradiance Reading of Irradiance Meter

An LED array emitting 455 nm wavelength of light, as verified by a calibrated spectrometer (41), was made to illuminate the uncalibrated irradiance meter as shown in Figure 3. The spectrometer equipped with a cosine corrector was placed adjacent to the irradiance meter such that they received the same intensity of light. Using the Ocean Insight OceanView Software Application (42), the absolute irradiance wizard was completed. Scans to average were set to 10 and boxcar width set to 10. The irradiance meter was set to output the raw sensor values to the serial line in the Arduino IDE. The pair of instruments were then placed 60 cm from the light source. The peak value measured by the spectrometer along with the raw data values output by the irradiance meter were both recorded at that distance for a total of 3 readings. The pair of devices were progressively moved towards the light source while true irradiance and raw output values were recorded. Irradiance vs raw sensor output was plotted and the linear relationship between the two was determined. The relationship was then implemented into the irradiance meter such that it would calculate the irradiance of the light source using only the sensor value. All the steps were repeated with the irradiance meter equipped with the AS7265x sensor and the relationship was plotted.

**Figure 3.**
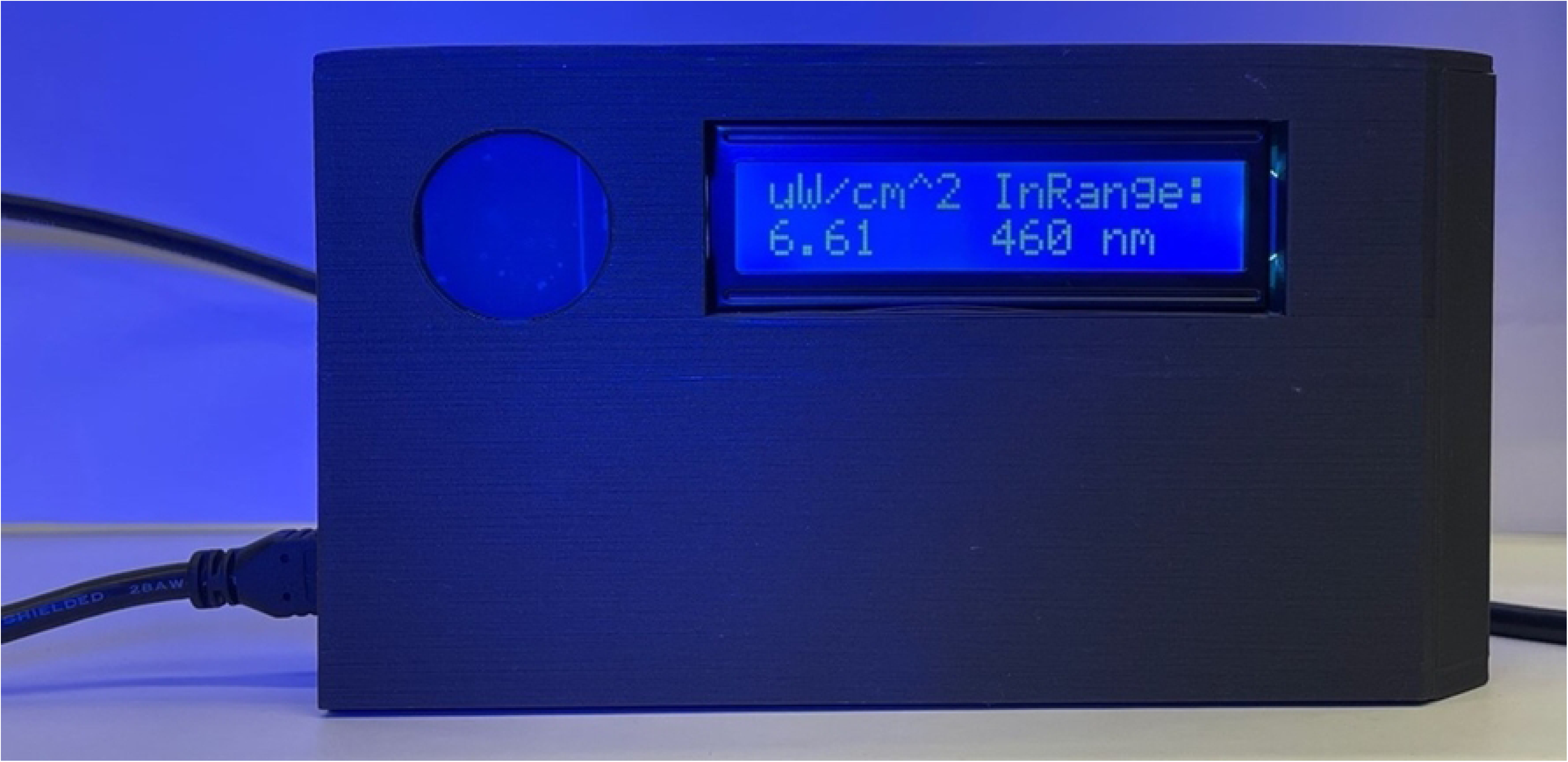
Line Diagram of Calibration Setup.

**Figure 4.**
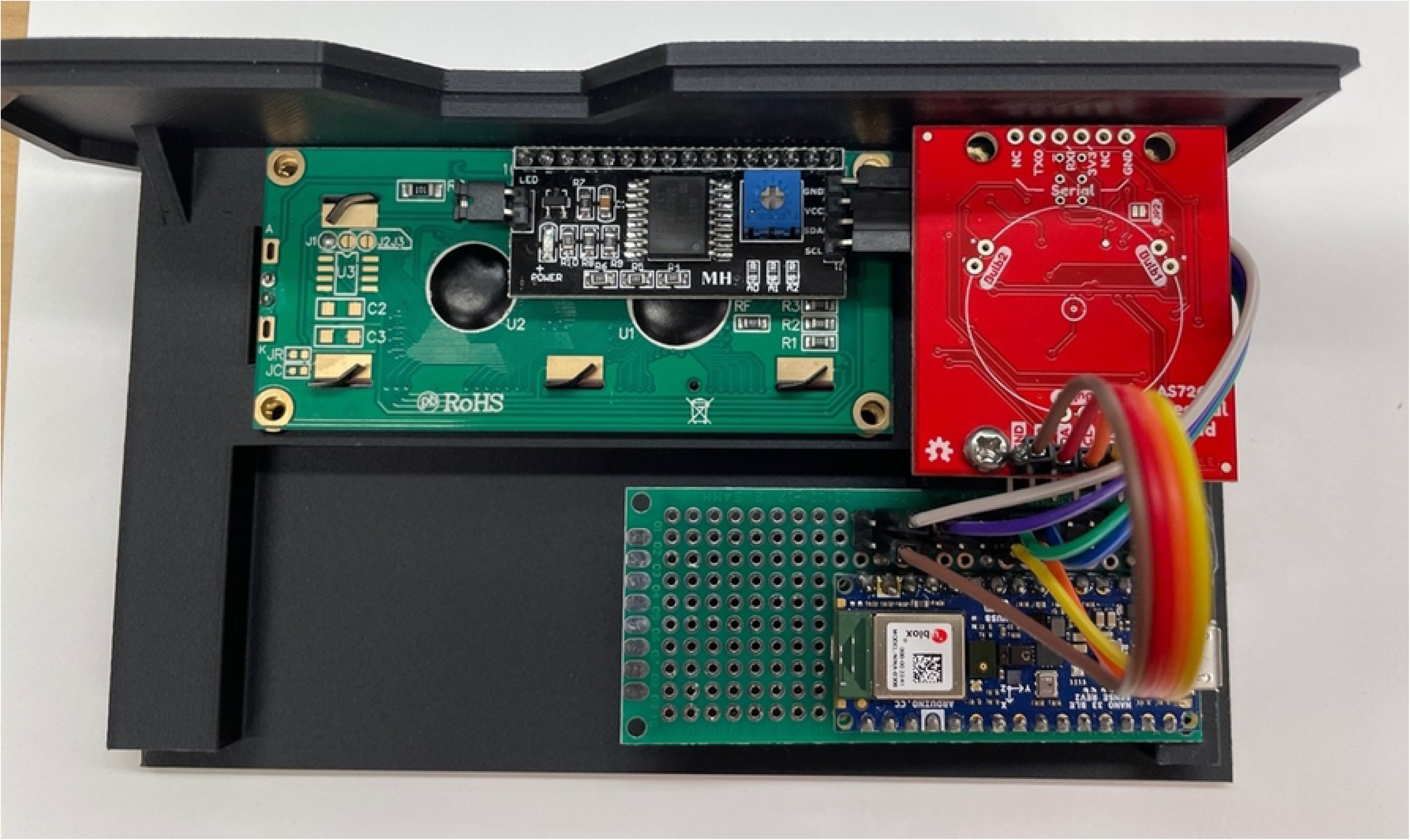
Calibration curve for ADPS – 9960-based open source system.

**Figure 5.**
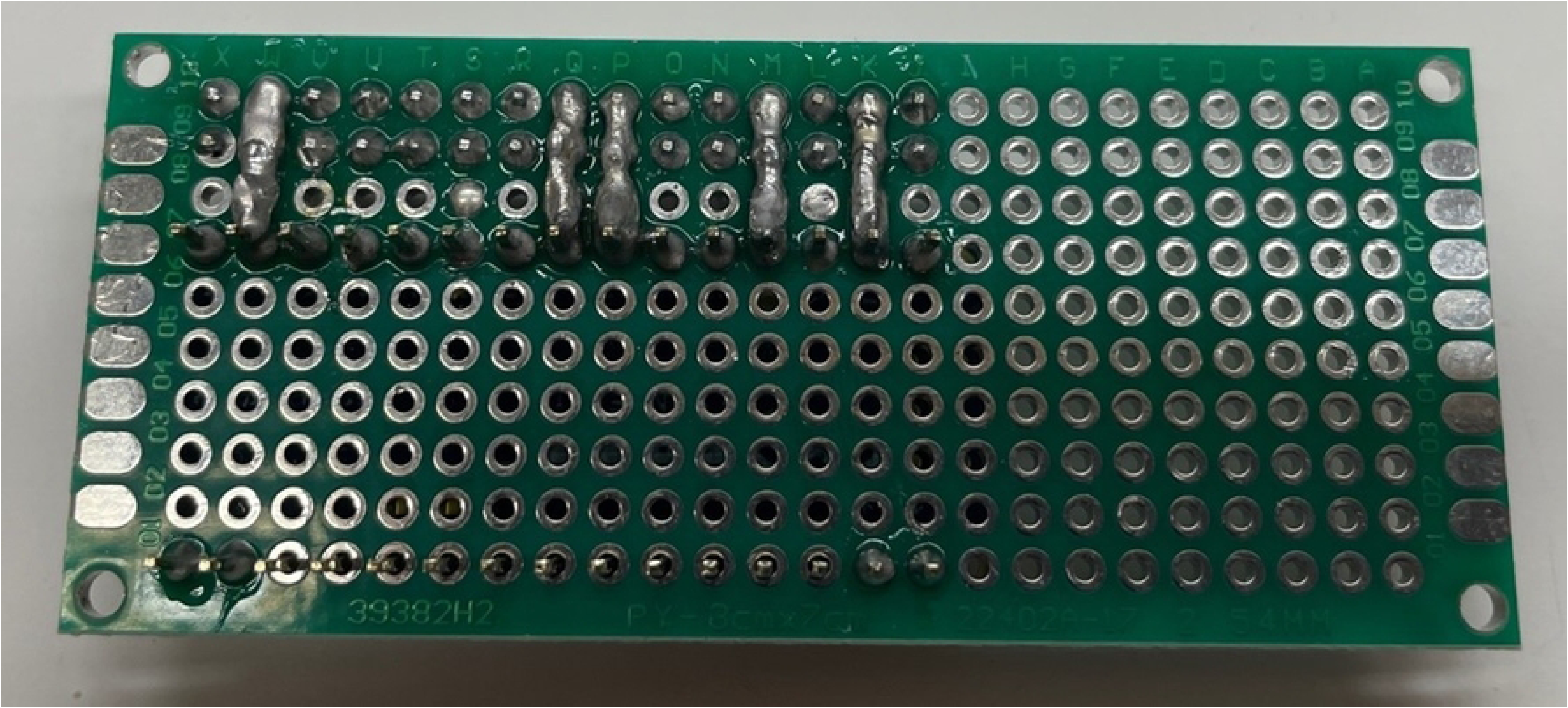
Calibration curve for AS7256x-based open source system.

### Calibrate Wavelength reading of Irradiance Meter

Depending on the sensor used in the build, the resolution of the wavelength reading differs. The built-in ADP-9960 sensor of the Arduino BLE Sense consists of 3 color channels: red – 625nm, blue – 465 nm and green – 525nm. The Sparkfun AS7265x consists of 18 color channel outputs ranging from 410 to 940 nm. First, a calibrated spectrometer was used to confirm the wavelength from the phototherapy light source. Both versions of the 3-D printed irradiance meter were then exposed to the same light and the output was observed.

### Testing Filter Spectral Shift

To create the filter, a tinted sheet of acrylic (43) with dimensions 25mm x 35mm x 5 mm was cut. A single layer of clear masking tape (44) was then placed on either side of the sheet. To determine the effect of the filter on the light source, a calibrated spectrometer was used to measure the spectrum of the 455 nm peak lamp source as the reference. The filter was then placed between the lamp source and the spectrometer. The spectrum was measured again and compared to the reference.

### Quantitative Testing

To determine accuracy, the light source was fixed at a point while spectral readings were taken at predefined locations of 10, 15, 20, 25, 30, 40, 50 and 60 cm from the source. The irradiance meter was then placed along those same locations and the values were compared against one another.

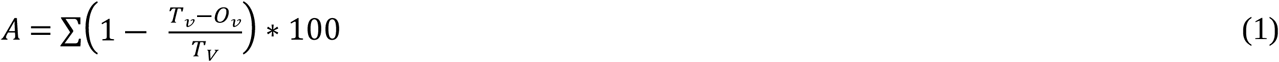

where A is accuracy, T_v_ is true value, O_v_ is observed value.

The precision of the device was determined by logging the device readings over a forty minute period while the light source was held at a constant intensity and distance. The variation in the output was measured to determine the device precision.

### Validation Tests in LMIC

A variable height test was performed between the open source light meter and an MTTS light meter (45). The MTTS light meter has an effective spectral response from 400-520nm, measurement range single 0.1 – 150.0 μW/cm²/nm and a resolution 0.1 μW/cm²/nm (45). The cosine characteristics of the MTTS light meter are ±2% at 30 degrees, ±7% at 60 degrees, and ±25% at 80 degrees with an accuracy or +/- 3% of reading. Tests were performed with a Photo-Therapy 4000 Jaundice Management machine from Drager (46).

## Results

### Irradiance Calibration Results

It was found that the ADPS9960 sensor had a linear relationship to the irradiance of the light source. The curve of best fit optimized by least squared regression with an R^2^ = 0.9947 showed the slope of the relationship as:

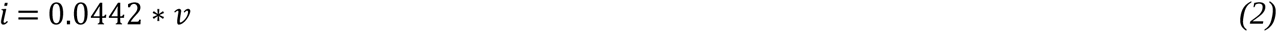

where i is irradiance and v is the raw value as shown in **Error! Reference source not found.**.

The AS7256x sensor also had a linear relationship to the irradiance of the light source. The curve of best fit optimized by least squared regression with an R^2^ = 0.9994 showed the slope of the relationship as:

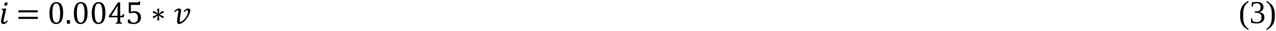

where i is the irradiance and v is the raw value as shown in **Error! Reference source not found.**.

### Wavelength Calibration Results

Figure 6 plots the spectral output of the phototherapy light to verify it falls within the 400 – 500 nm range. Its peak was found to be at 455 nm. Because the ADPS 9960 sensor is limited to three color channels at 465, 525 and 625 nm. It can determine that the highest peak is within 465 525 nm but cannot identify the precise wavelength nor can it identify a peak with a wavelength below 465 nm with any certainty.

**Figure 6.**
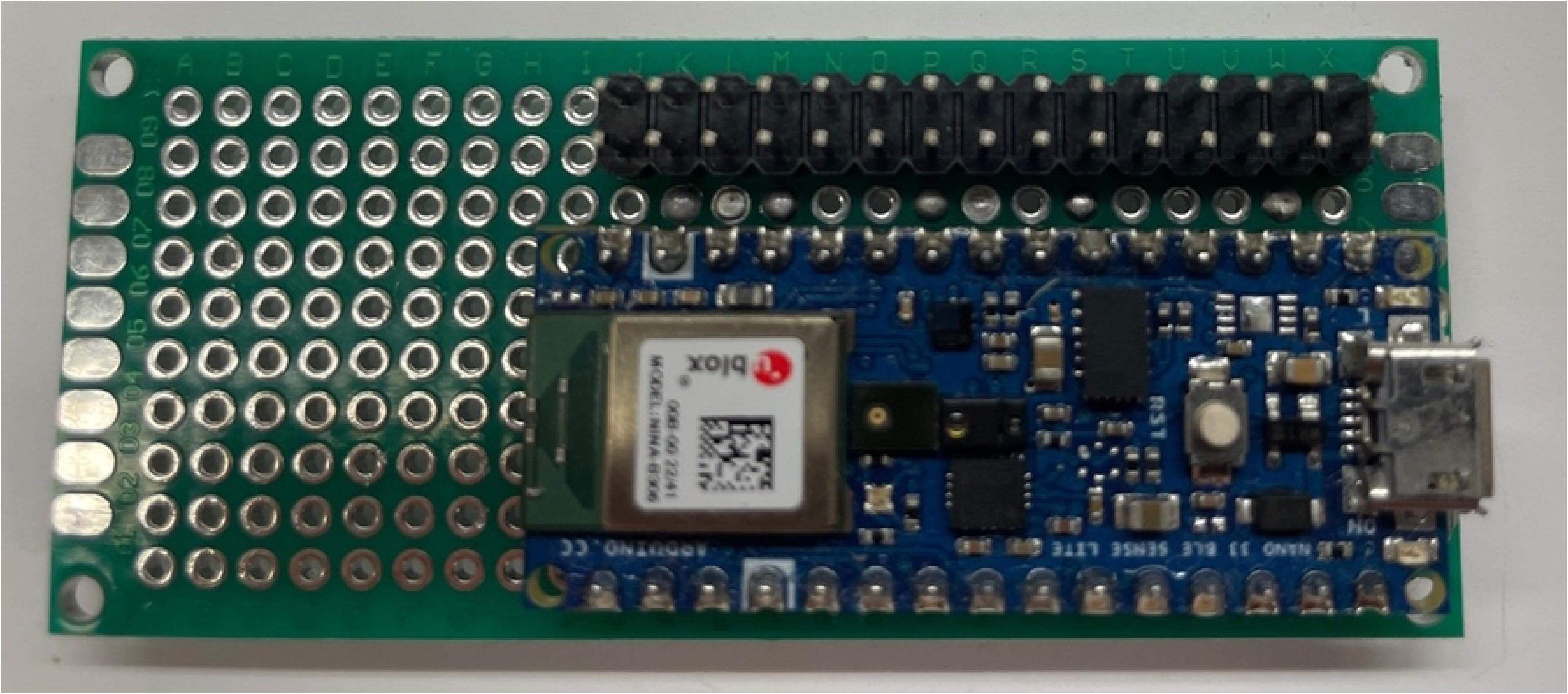
Spectral Analysis of Phototherapy Source

The device using the AS7256x sensor was programed to indicate that the light source was within range if the largest peak was found in the 435nm, 460nm, or 485 nm channel. Figure 7 shows the output of the AS7256x equip device when exposed to the phototherapy source. The AS7265x sensor could verify that the detected wavelength was between 410 – 485 ± 12.5 nm.

**Figure 7.**
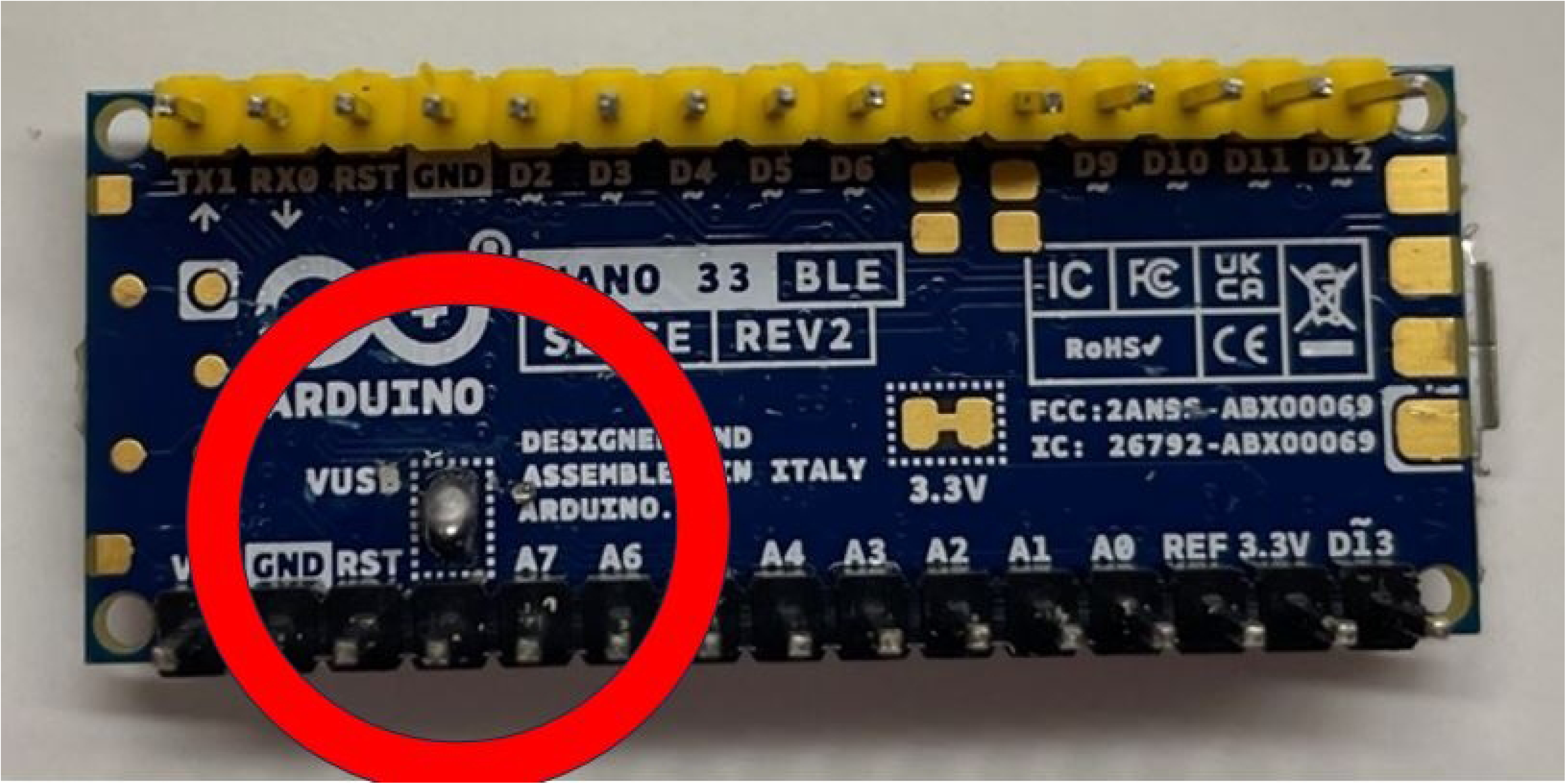
AS2756x Irradiance Meter Displaying Light Source Characteristics

The APDS-9960 version did not meet the requirement by the UNICEF Supply Catalogue (21) for detection within 400 - 500 nm range since wavelengths below 465 are not accurately recorded. The AS7256x meets the requirement by the UNICEF Supply Catalogue (21) for detection within 400 500 nm range.

### Results of Spectral Shift

The results of the filter test are shown in Figure 8. As can be seen, the peak of both the source and the filtered light occurs at 455 nm therefore, the filter does not cause any spectral shift in the 455 nm range. Secondly, the peak irradiance of the spectrum with the filter is significantly lower. Thus, the intended effect of the filter has been achieved. The maximum exposure of the sensor has been lowered such that it will operate within the desired range and the wavelength of light was verified to not shift as a consequence of passing through said filter.

**Figure 8.**
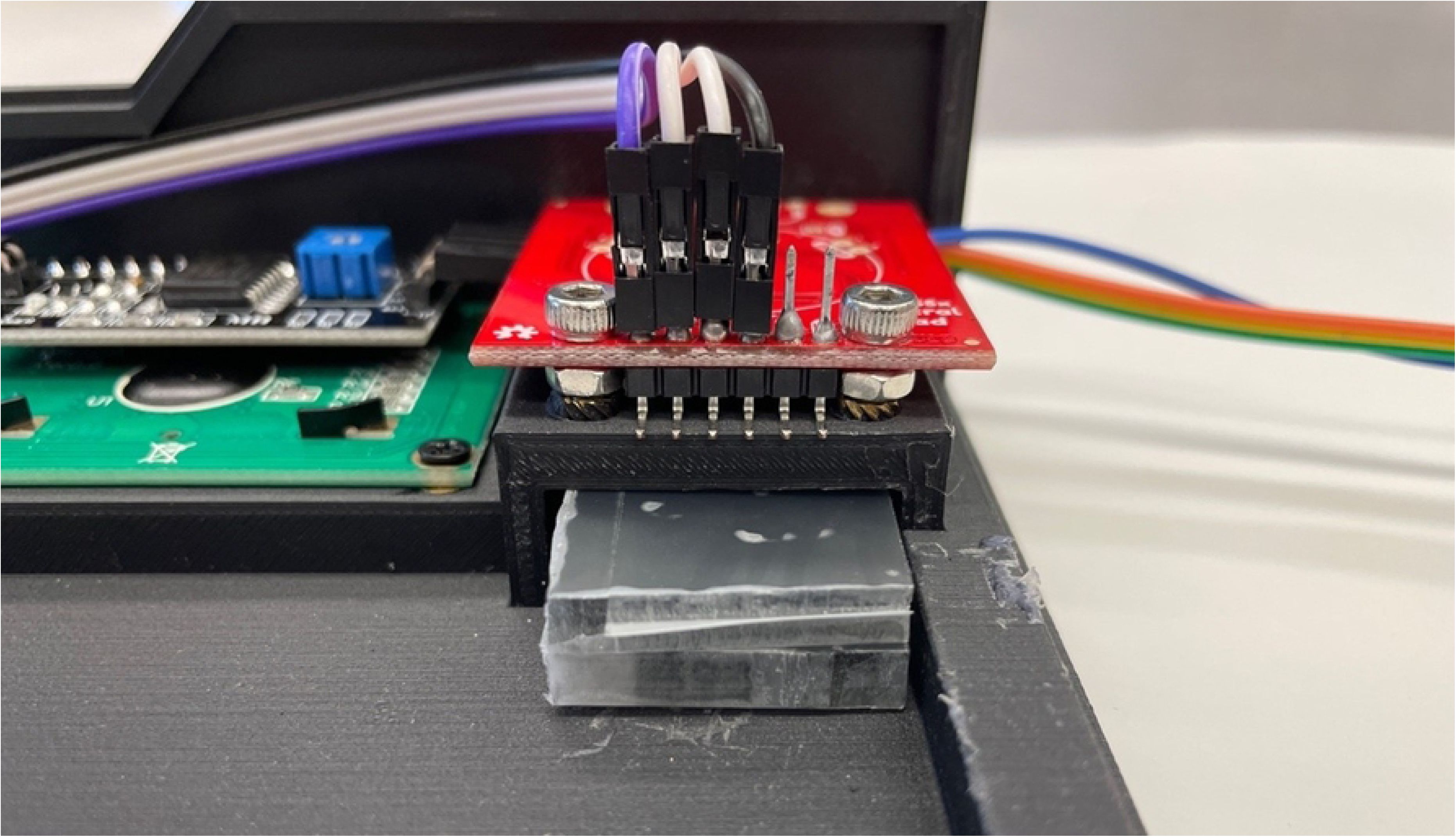
Effect of Filter on Phototherapy light.

### Irradiance Accuracy

The true value compared with that of the AS2756X sensor can be observed in Table 2. The final accuracy is 98.64% using **Error! Reference source not found.**.

**Table 2.** Comparison of true value vs AS2756X irradiance reading.

| Distance (cm) | True Value (uW/cm<sup>2</sup>/nm) | Irradiance meter reading (uW/cm<sup>2</sup>/nm) | Delta | Accuracy |
| --- | --- | --- | --- | --- |
| 60 | 22.5 | 22.37 | 0.13 | 0.9942 |
| 50 | 32.17 | 32.42 | 0.25 | 0.9922 |
| 40 | 48.32 | 49.73 | 1.41 | 0.9708 |
| 30 | 79.88 | 80.15 | 0.27 | 0.9966 |
| 25 | 104.36 | 104.43 | 0.07 | 0.9993 |
| 20 | 145.33 | 143.12 | 2.21 | 0.9848 |
| 15 | 208.32 | 201.35 | 6.97 | 0.9665 |

Figure 9 plots the irradiance meter readings over 40 minutes while exposed to a fixed light source. The AS2756x irradiance meter outputs an average value of 21.74 ± 0.04 uW/cm^2^/nm.

**Figure 9.**
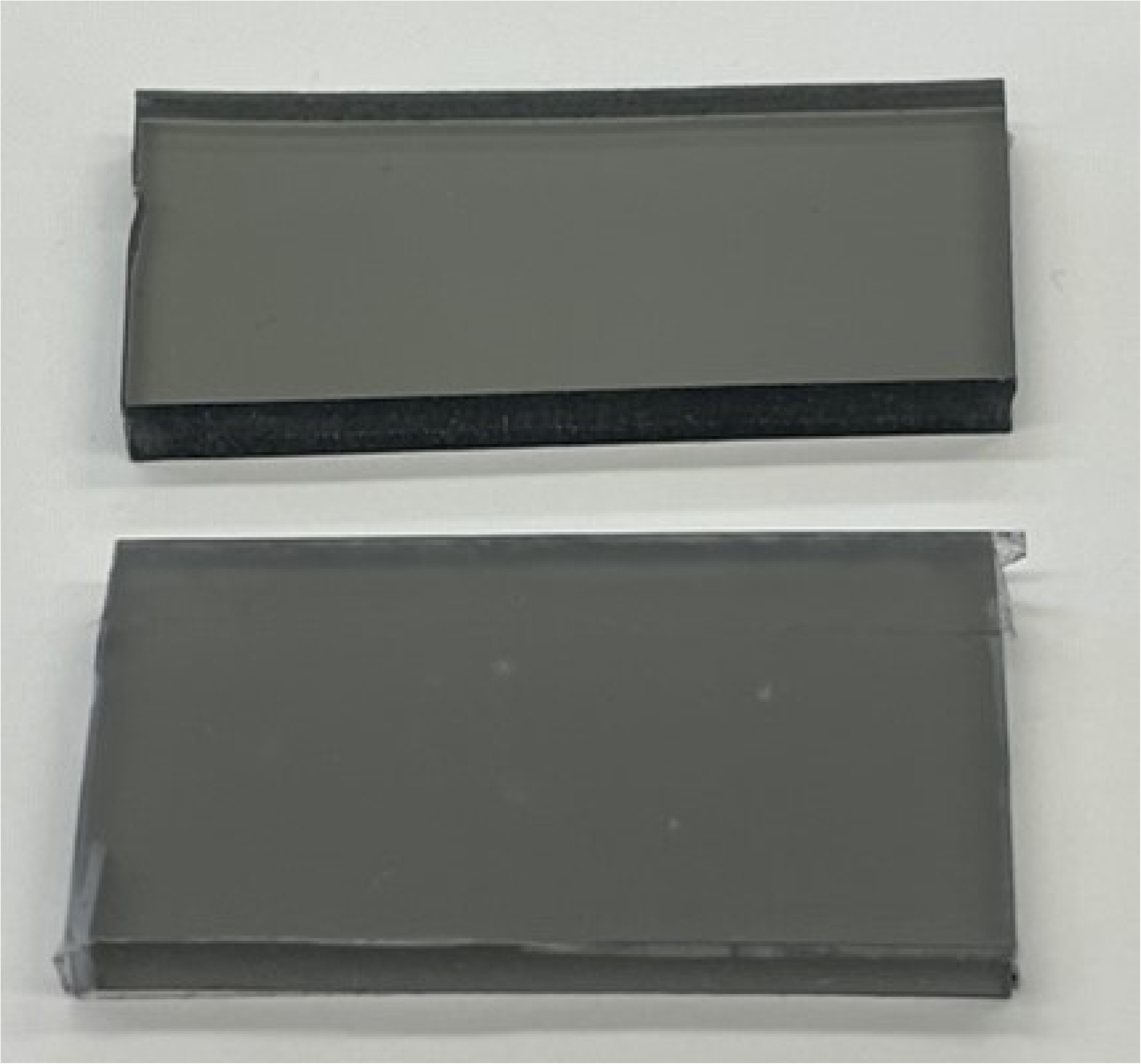
AS2756X Irradiance meter under constant light Intensity for 1 hour.

The true value compared with that of the ADPS-9960 sensor can be observed in Table 3. The final accuracy is 90.40% using **Error! Reference source not found.**.

**Table 3.** Comparison of true value vs ADPS9960 irradiance reading.

| Distance (cm) | True Value (uW/cm2/nm) | Irradiance meter reading (uW/cm2/nm) | Delta | Accuracy |
| --- | --- | --- | --- | --- |
| 60 | 17.21 | 19.45 | 2.24 | 0.8698 |
| 50 | 23.84 | 27.18 | 3.34 | 0.8599 |
| 40 | 36.78 | 39.99 | 3.21 | 0.9127 |
| 30 | 58.2 | 63.95 | 5.75 | 0.9012 |
| 25 | 78.55 | 84.17 | 5.62 | 0.9285 |
| 20 | 109 | 117.59 | 8.59 | 0.9212 |
| 15 | 154.68 | 164.8 | 10.12 | 0.9346 |
| 10 | 242.78 | 243.17 | 0.39 | 0.9984 |

Figure 10 plots the irradiance meter readings over 40 minutes while exposed to a fixed light source. The AS2756x irradiance meter outputs an average value of 32.92 ± 0.71 uW/cm^2^/nm.

**Figure 10.**
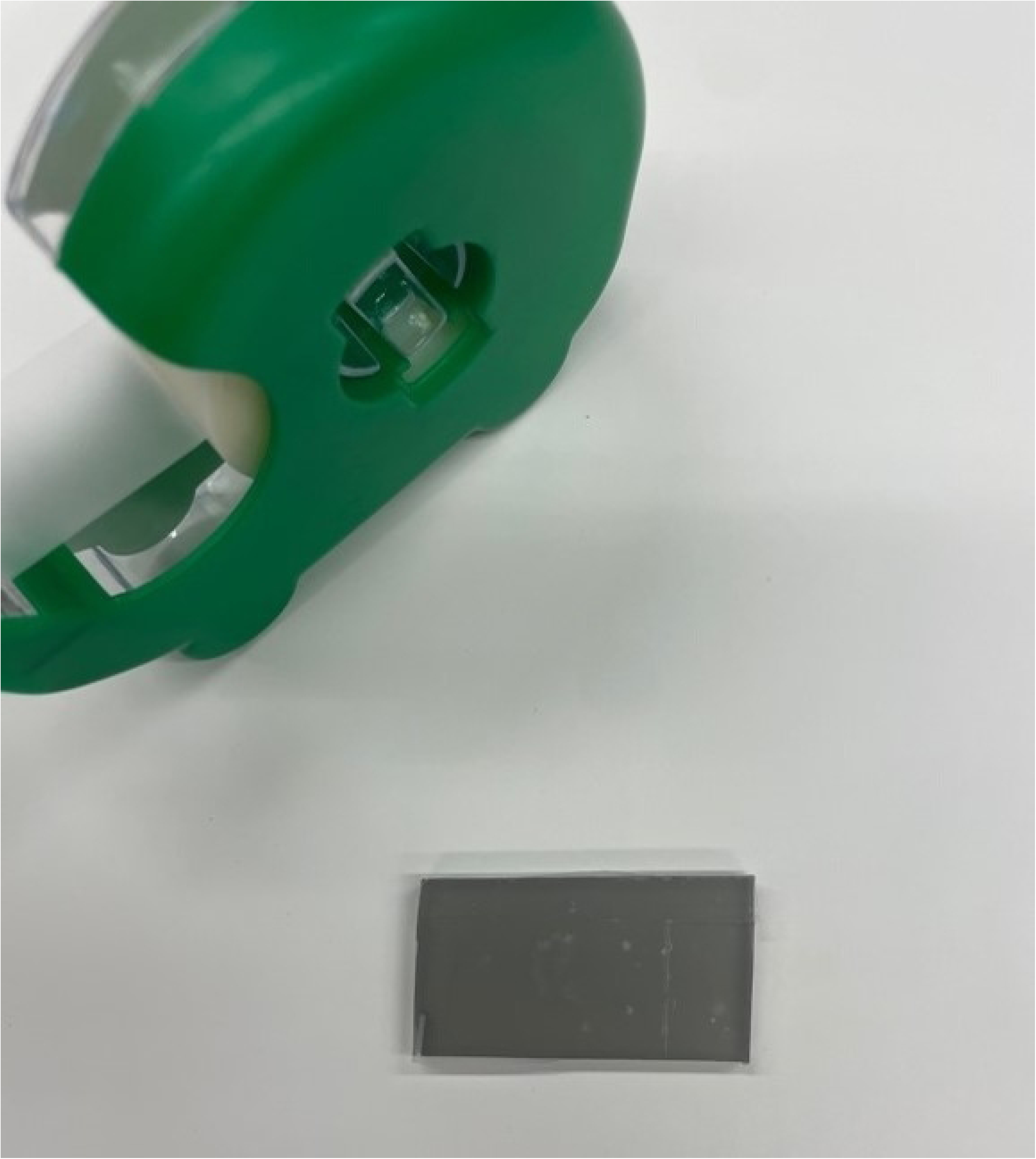
ADPS9960 Irradiance meter under constant light intensity for 1 hour.

### Validating Results in LMIC –

The validation results on the Photo-Therapy 4000 Jaundice Management machine from Drager in an LMIC are shown in Table 4.

**Table 4.** Light meter responses as a function of distance from a Photo-Therapy 4000 Jaundice Management machine.

| Distance from light | OS light meter | MTTS | Error |
| --- | --- | --- | --- |
| 20 | 18.4 | 21 | 0.124 |
| 25 | 36.6 | 36 | 0.017 |
| 30 | 45.05 | 42 | 0.073 |
| 40 | 50.05 | 51 | 0.019 |
| 50 | 68.18 | 69 | 0.012 |
| 60 | 79.3 | 79 | 0.004 |

**Table 4.** Print Settings

|  |  |
| --- | --- |
| Nozzle Temperature | 210 C |
| Bed Temperature | 60 C |
| Infill | 15% |
| Supports | none |
| Layer Height | 0.2 mm |
| Perimeters | 3 |
| Speed | 40 mm/s |
| First layer speed | 20 mm/s |
| Brim | not required |

As can be seen by the results in Table 4, the results of the open source light meter are in good agreement with the commercial MTTS system at larger distances that would be clinically relevant. The greater errors observed at small distances are due to cosine errors and expected.

## Discussion

This paper has demonstrated that a 3-D printed irradiance meter using the AS2756x sensor is an effective device to calibrate phototherapy light sources. Irradiance can be measured up to 200 uW/cm^2^/nm, surpassing the UNICEF requirements (47), with an accuracy of 98.6% and will determine the wavelength of peak spectrum with an accuracy of ± 12.5 nm. Over a forty minute period the device is stable within ± 0.04 uw/cm^2^/nm. The additional cost of the AS7265x sensor, however, may be a consideration for use cases in LMIC.

The APDS-9960 sensor meets the spectral irradiance range requirements of 150 uW/cm^2^/nm. Compared to the AS2756X sensor, the ADPS-9960 is limited in its ability to verify wavelength and provides less accurate readings of irradiance intensity. The APDS-9960 only validates wavelengths between 465 – 525 nm so does not meet the wavelength requirements set out by the UNICEF supply datasheet (47). Given the lower accuracy, lower precision and limited wavelength reading, the ADPS- 9960 version is not generally recommended (14). In highly resource constrained settings the ADPS- 9960 version may be considered with the understanding that the device is rated only for LED sources with a peak above 465nm.

As previous work has shown, effective treatment of hyperbilirubinemia is commonly administered through light therapy of 400 – 500 nm with an intensity greater than 30uW/cm^2^/nm. The light source was verified by a calibrated spectrometer (Figure 6) providing consistency between previously published work and the work shown here.

## Limitations and Future Work

Consideration should be made for the long-term performance of the device. The PLA housing of the device is not expected to degrade as long as the device is protected from outside elements (48).Voltage regulation is managed on board the Arduino nano while additional regulation is provided in the battery bank of models using a portable battery (Appendix A3). Future work may focus on the long-term stability of the device and over what period recalibration of the unit is required.

## Conclusions

It has been demonstrated that for under $200 CAD, one can construct a phototherapy calibration device capable of measuring up to 200uW/cm^2^/nm with an accuracy of 98.6% and detect the peak wavelength within +/- 12.5 nm. 3D printed open-source irradiance meters are a viable option for calibrating phototherapy units in LMIC to treat hyperbilirubinemia.

## Data Availability

Files, code and data can be found at https://osf.io/7dqp6/ DOI 10.17605/OSF.IO/7DQP6

https://doi.org/10.17605/OSF.IO/7DQP6

## Acknowledgments

This work was supported by the Thompson Endowment and Frugal Biomed Catalyst Grant. The authors would like to thank C. Brooks and G. Antonini for helpful discussions.

Files, code and data can be found at https://osf.io/7dqp6/DOI10.17605/OSF.IO/7DQP6

## Appendix

### A1: Build Instructions for Light Calibration Box (ADPS 9960 SENSOR)

Download and 3-D print the case in any RepRap-class (49–51) fused filament 3-D printer oriented as shown in Figure 11 with the input parameters show in Table 5.

**Figure 11.**
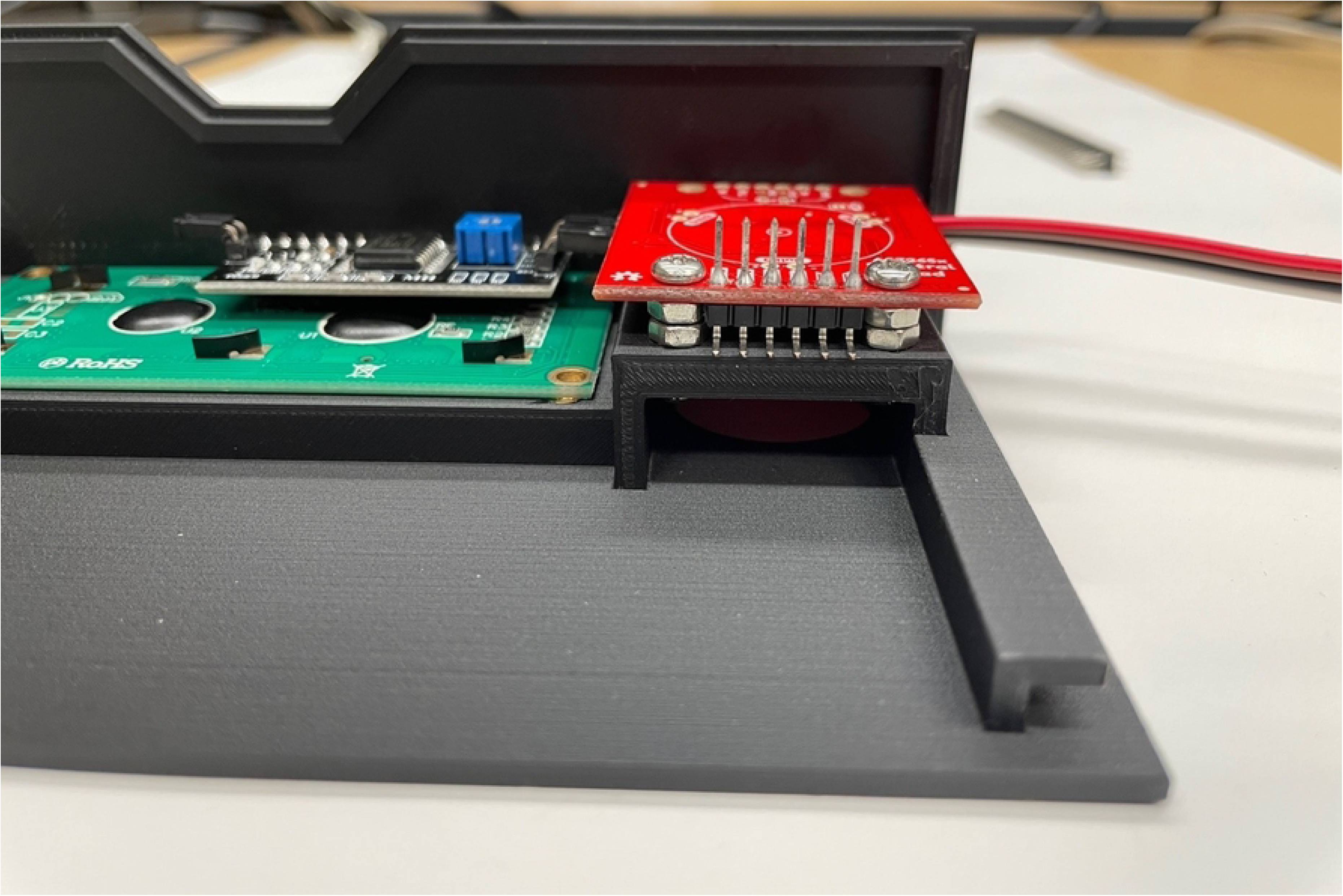
Print Orientation for ADPS 9960 Case

**Table 5.** 3-D Print Settings

|  |  |
| --- | --- |
| Nozzle Temperature | 210 C |
| Bed Temperature | 60 C |
| Infill | 15% |
| Supports | recommended |
| Layer Height | 0.2 mm |
| Perimeters | 3 |
| Speed | 40 mm/s |
| First layer speed | 20 mm/s |
| Brim | not required |

1. Gather the parts as shown or see bill of materials.
2. Install heat inserts with a hot soldering iron.
3. Connect the I2C adpater to the LCD screen.
4. Place a single layer of clear tape of each side of the plexi glass filter.
5. Slot the filter into place and secure screen assembly to case with M3 screws.
6. The LCD Screen only works with 5V power. To access 5V from the pins, solder the VUSB pads together as shown.
7. Connect the Arduino and screen together. GND pin goes to Arduino GND, VCC connects to Arduino 5V, SDA connects to pin 4 and, SCL connects to pin 5. Hot glue can then be placed on the rails which supports the microcontroller.
8. The Arduino is secured in place as shown until the glue solidifies. Note: the orientation of the Arduino can be flipped depending on whether you would like the USB port to face inside or outside the case. Facing the port inwards allows more convenient battery-operated access.
9. Upload the ‘<u>ADPS9960_LightCalibration.ino’</u> file to the board and close case. Power from USB port to operate.

### Appendix A2. Build Instructions- Light Calibration Box with SPARKFUN AS7265x Sensor

Download and 3-D print the case in any RepRap-class (49–51) fused filament fabrication (FFF) 3-D printer with the input parameters show in Table 5.

1. Gather the required components as per BOM (see Appendix A).
2. Install heat inserts using the soldering iron as shown in Figure 22.
3. Solder the LCD screen too I2C Adapter as shown and secure to the case with M3 screws as shown in Figure 23.
4. Screw down the AS7265 Sensor using either a standoff or a pair of hex nuts as a spacer.
5. Remove the protective film from the plexiglass filter and apply a single layer of masking tape to both the top and bottom face of the filter. Insert and hot glue the filter in the filter slot. The clear tape is required to further attenuate the incoming light so that the filter operates within range.
6. For the Arduino nano to provide 5V, the VUSB pads need to be soldered together as shown.
7. The Arduino can then be soldered to the protoboard along with the pin headers as shown. Note: the usb connector should extend beyond the board as shown with the pin headers closer to the analogue pin side (left side) of the board.
8. 8) Solder the ground pin, 5V pin, 3V pin, A4 pin and A5 pin across to their corresponding headers as shown.
9. Wire the LCD and AS2656x sensor ground wires to ground on the Arduino. Wire the 5V output pin of the Arduino to the LCD Screen 5V input. Wire the 3V output pin of the Arduino to the 3V input of the ADPS sensor. Both the screen and the sensor have SDA and SCL pins that should be connected to pins 4 and pins 5 respectively.
10. Upload code from either source the OSF: https://osf.io/7dqp6/ or

GitHub: https://github.com/JoshuaGivans-uwo/FAST_IrradianceMeter

1. 10. Close Case
2. 11. Power though battery bank or any other 5V DC source (PC, wall adapter ect.). Consider printing Case_File_Large.stl if a large sized power bank is used,
3. 12. Ocean Insight, calibrated light, and spectrometer Wavelength, irradiance, distance from source
4. 13. Calibration curve from Ocean Optics and how you use the software Figure showing peak irradiance at 465 nm at specific distance from light source Possible files to include:

Calibration curve for cosine corrected fiber optic STL Files for case

### Appendix A3. Build Instructions – Battery Option

Either device may be powered from a mobile power bank. Any power bank supplying at least 5V will work. The specific power bank used in this version can be found in the BOM. The larger case which was designed to house this particular bank can be found as an STL file in the OSF repository (52).

NOTE: Most power banks have a low power detection system which powers off when the current drawn is too low. This system is triggered when connected to the Arduino and will automatically power down after 20 seconds. A double tap on the power button after it is on will switch the battery bank into low power mode to avoid automatic shut off. It is important that this feature is available if another power bank is used.

Assembly is the same as Appendix A2 with the additional step that the battery is tapped to the inside of the back of the unit. If the battery bank used is larger than 15.75 x 7.37 x 1.91 cm it is recommended to print the original case and mount the battery on the outside of the device.

**Figure 12.**
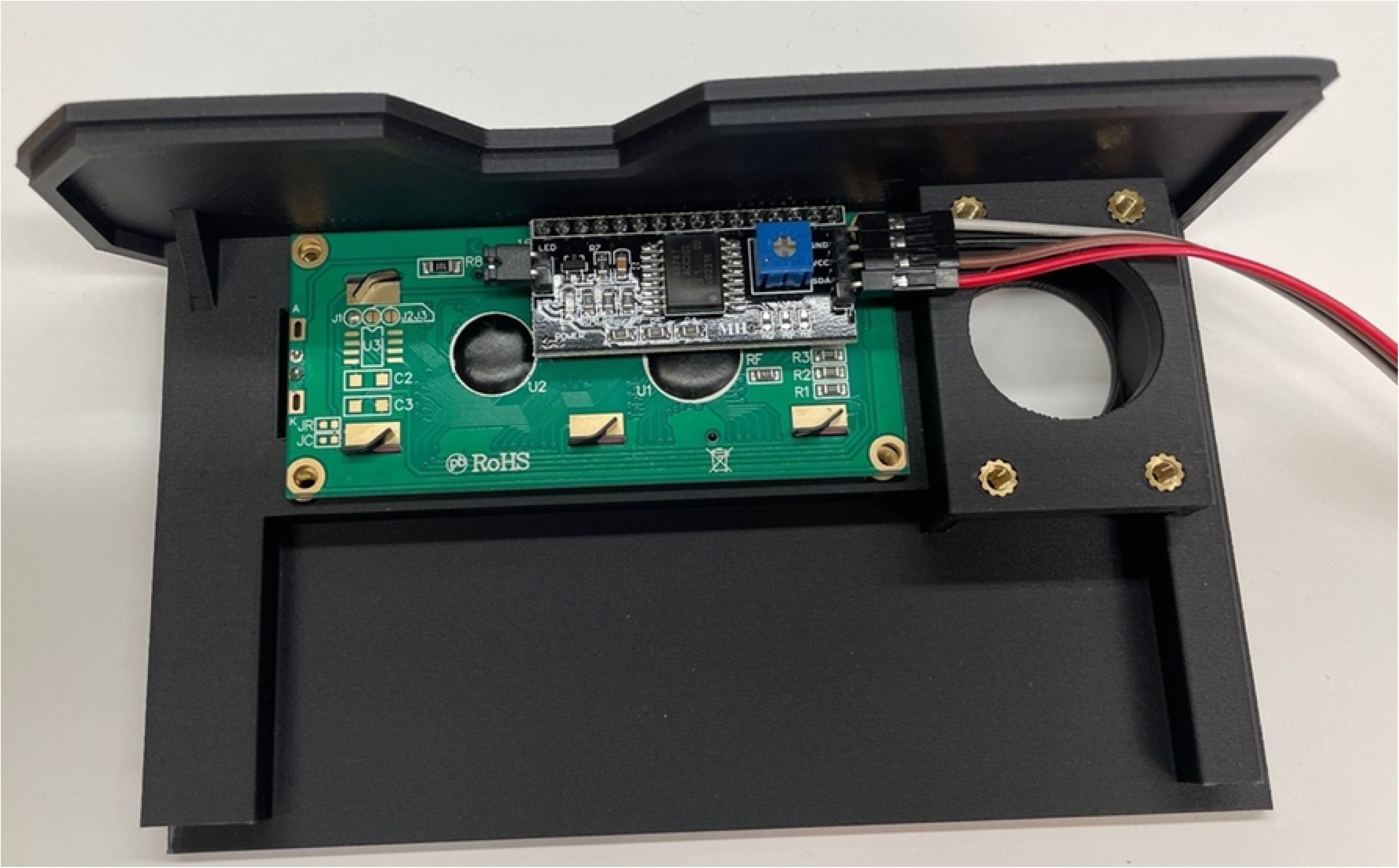
Gathered components.

**Figure 13.**
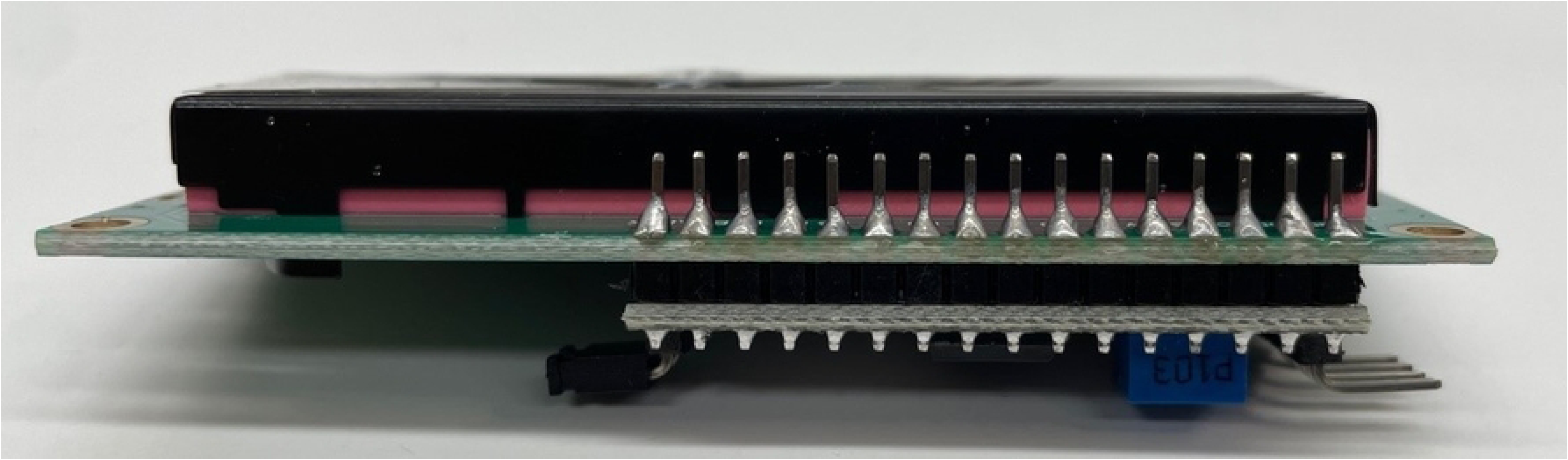
Heat inserts in place.

**Figure 14.**
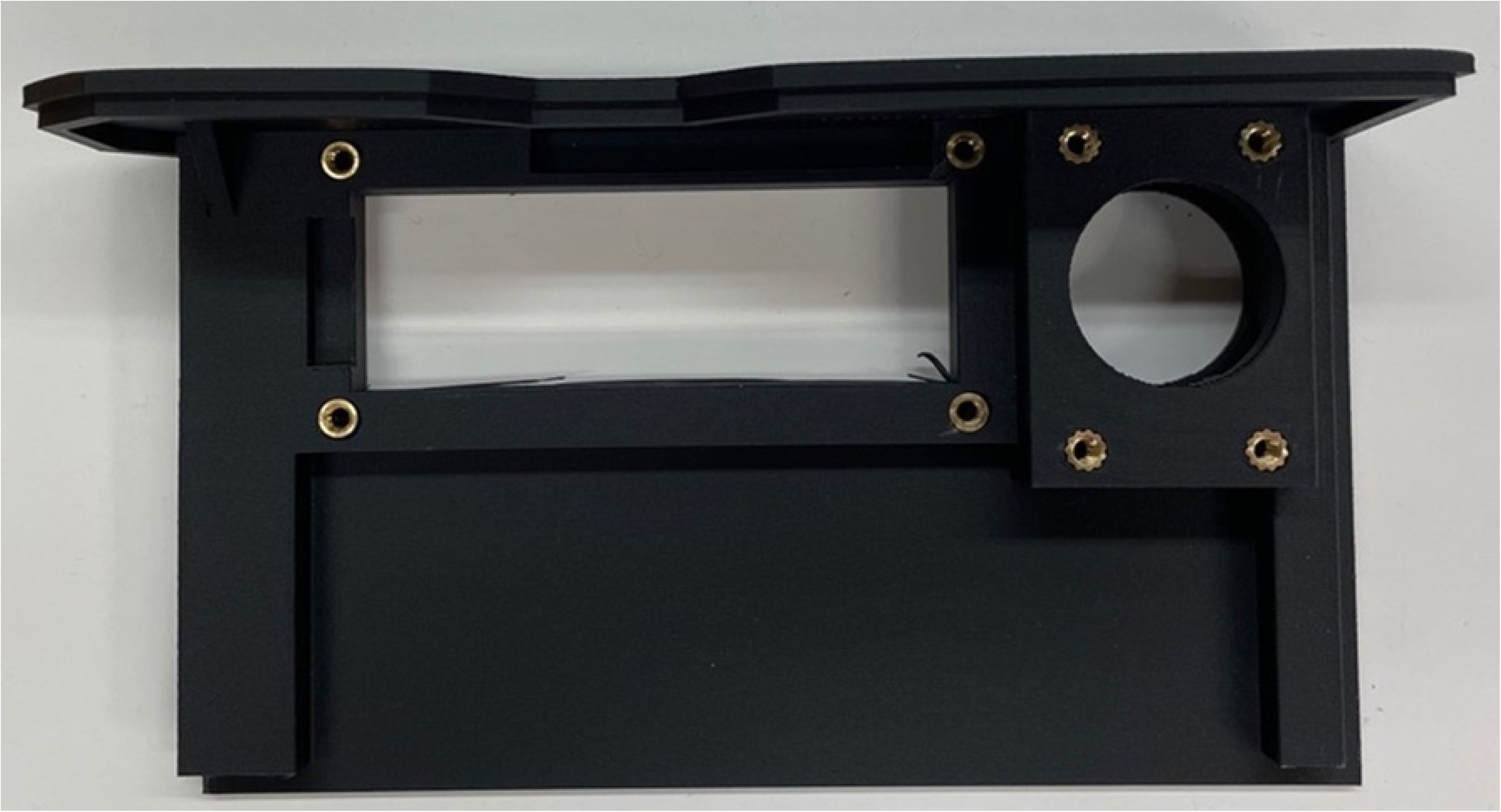
I2C LCD Assembly

**Figure 15.**
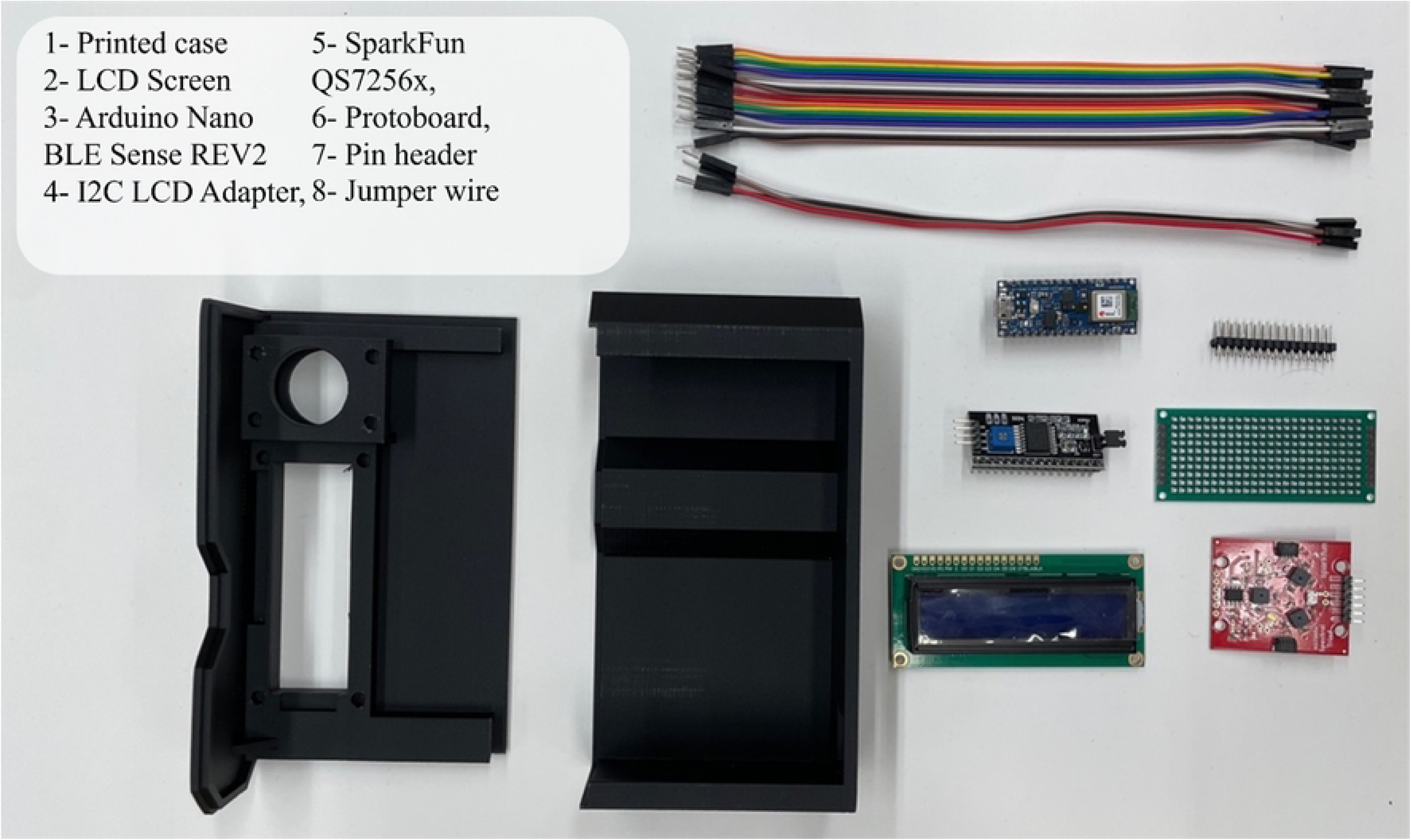
Filter preparation.

**Figure 16.**
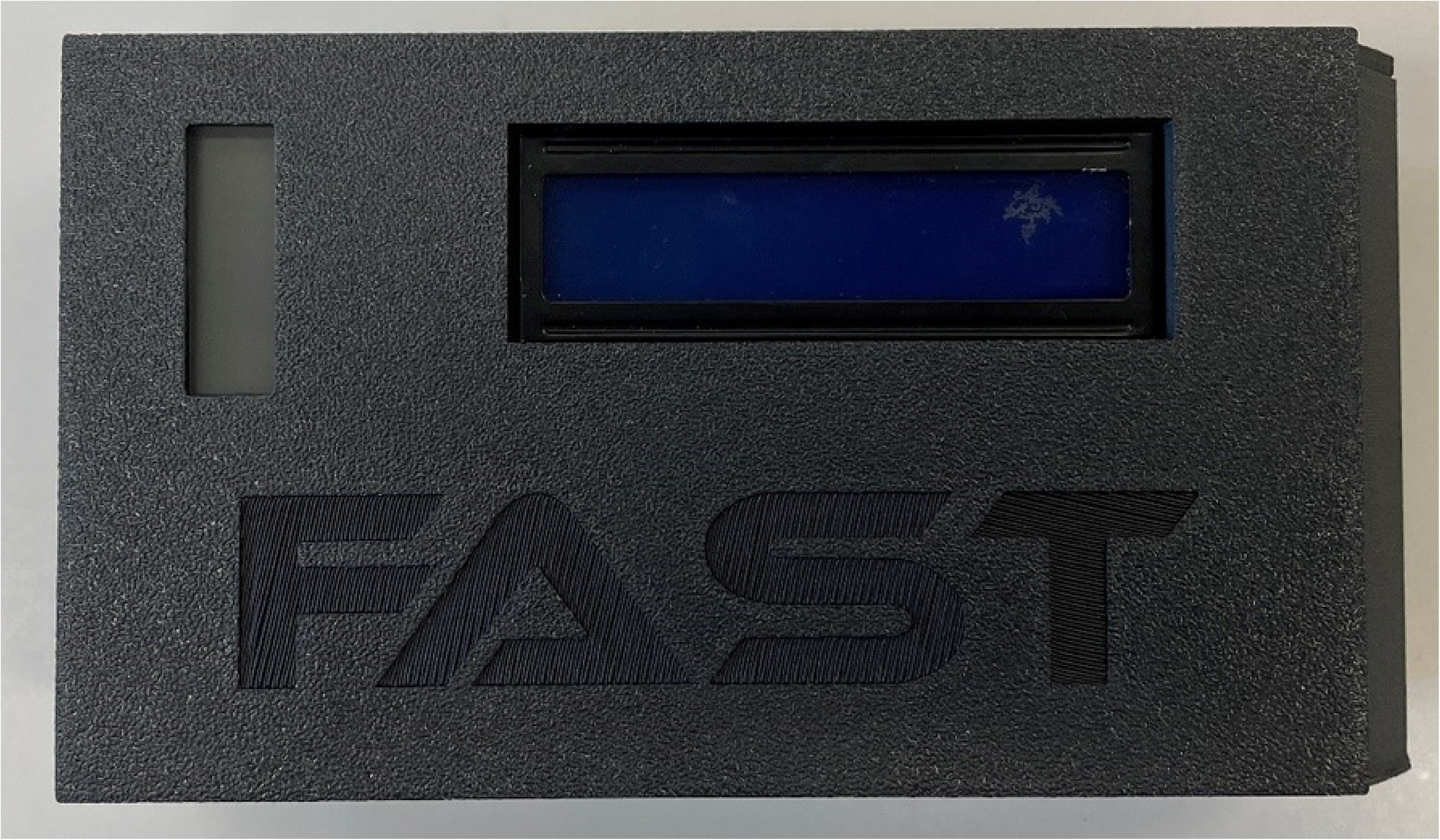
Screen and Filter in place.

**Figure 17.**
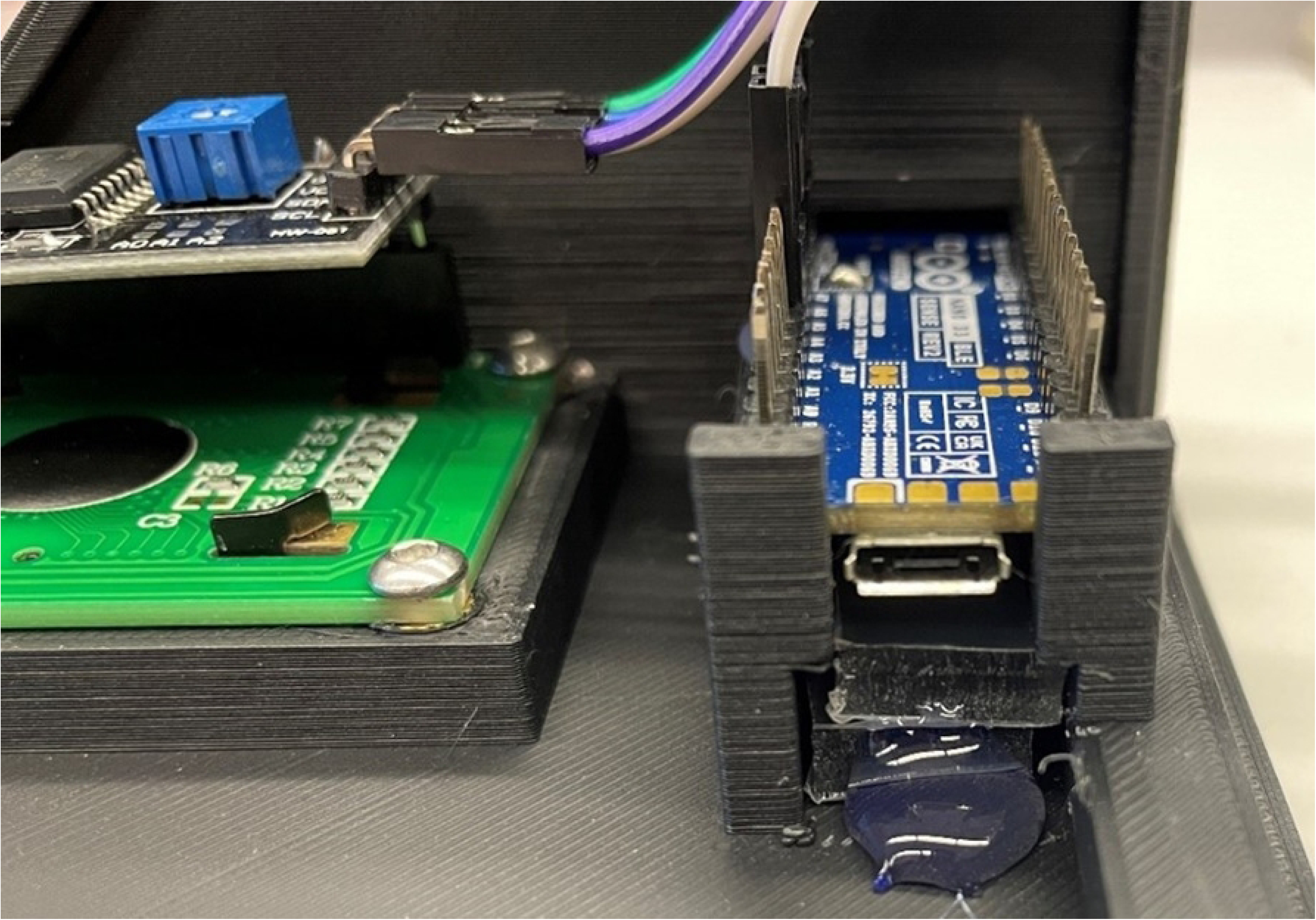
Soldered pads to access 5V.

**Figure 18.**
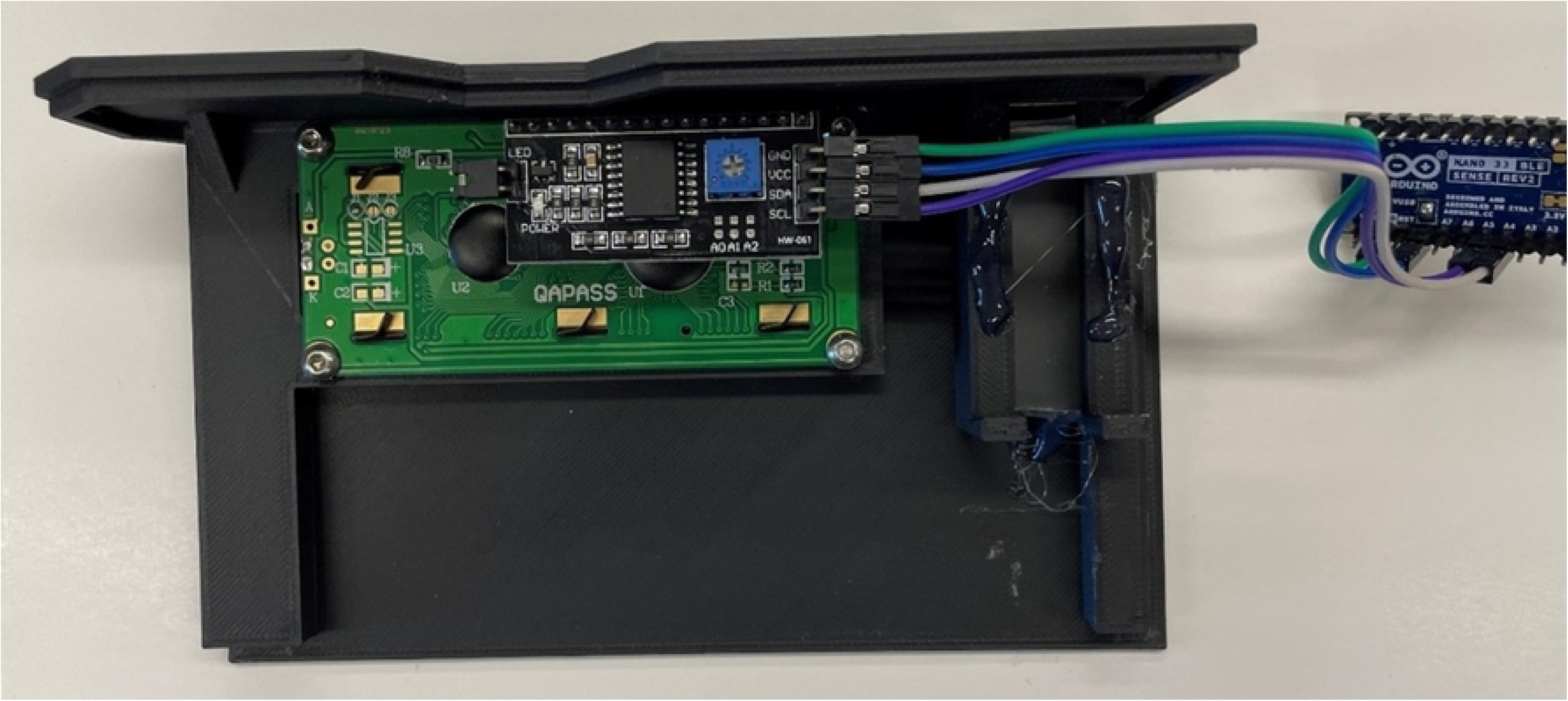
Screen connected to Arduino microcontroller.

**Figure 19.**
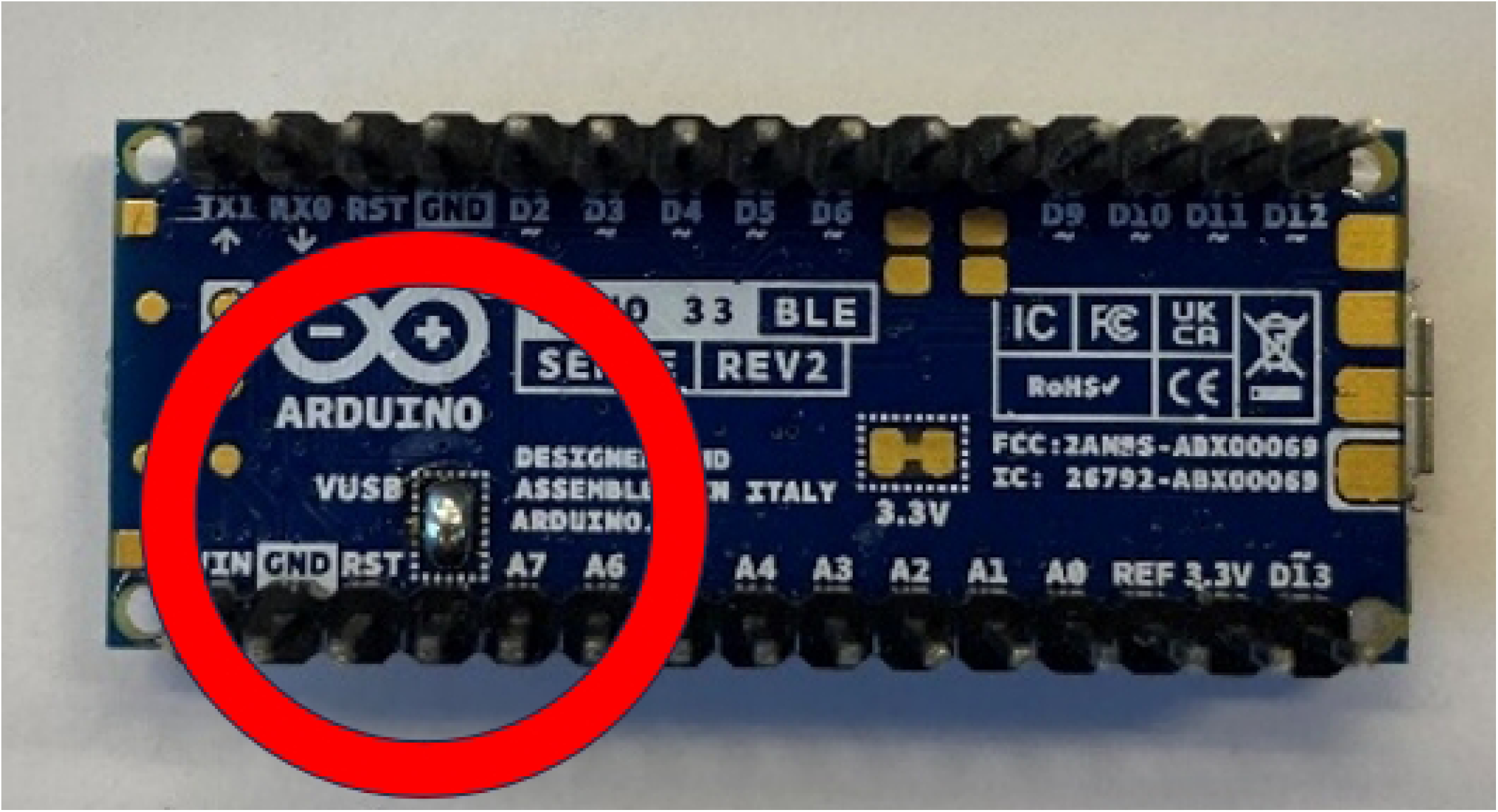
Arduino mounted in place.

**Figure 20.**
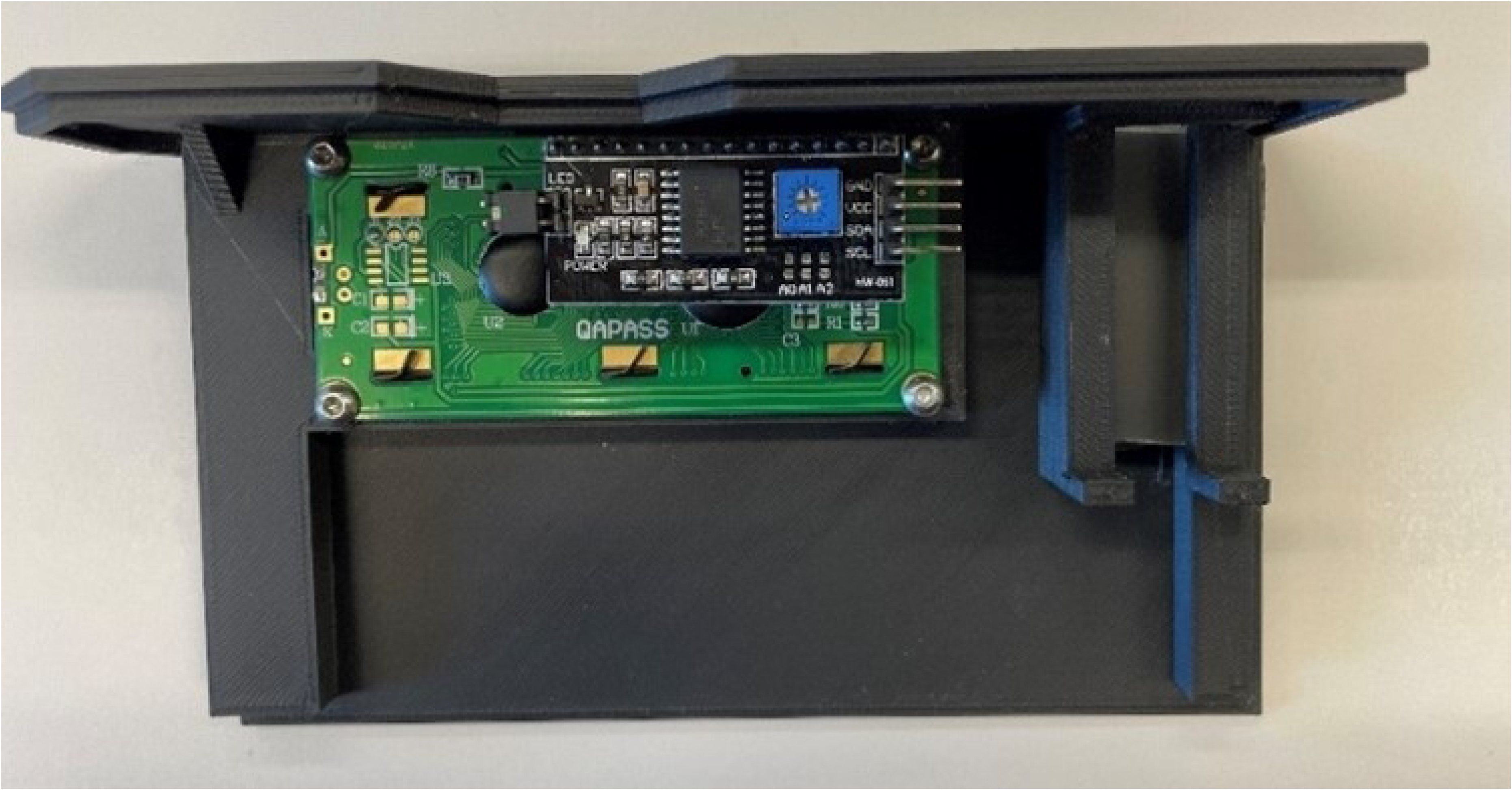
Completed Irradiance Meter with ADPS9960 Sensor.

**Figure 21.**
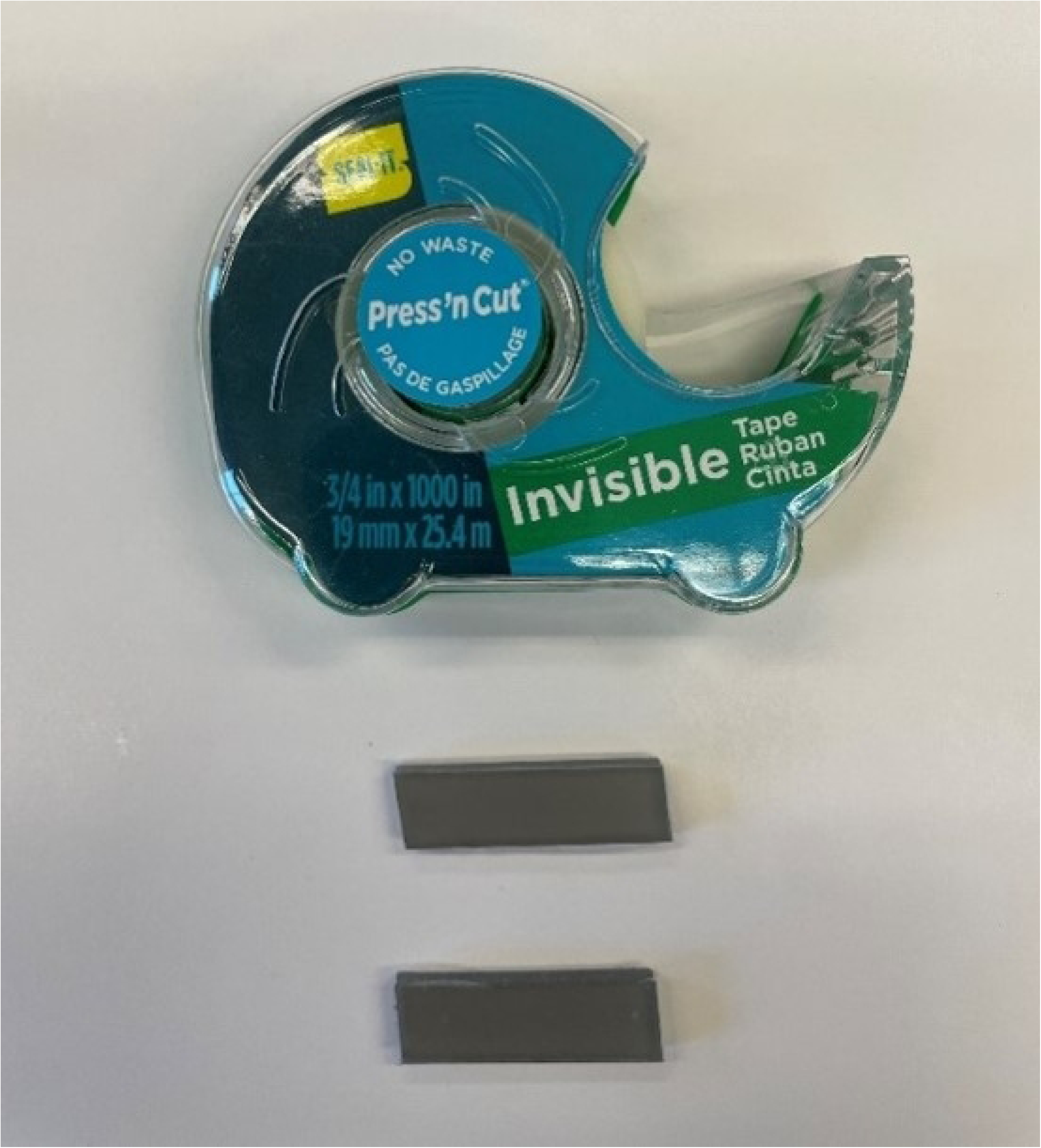
Components of SparkFun AS7265x sensor-based open source light calibration box.

**Figure 22.**
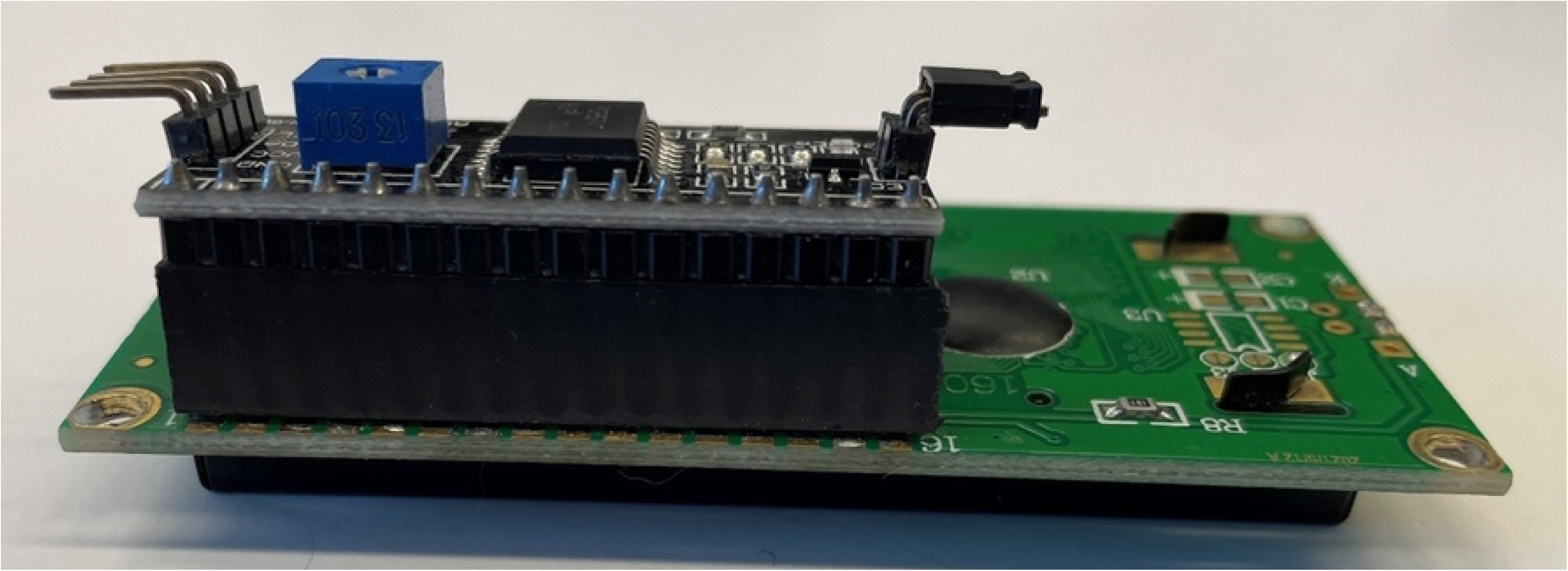
Device with heat inserts installed.

**Figure 23.**
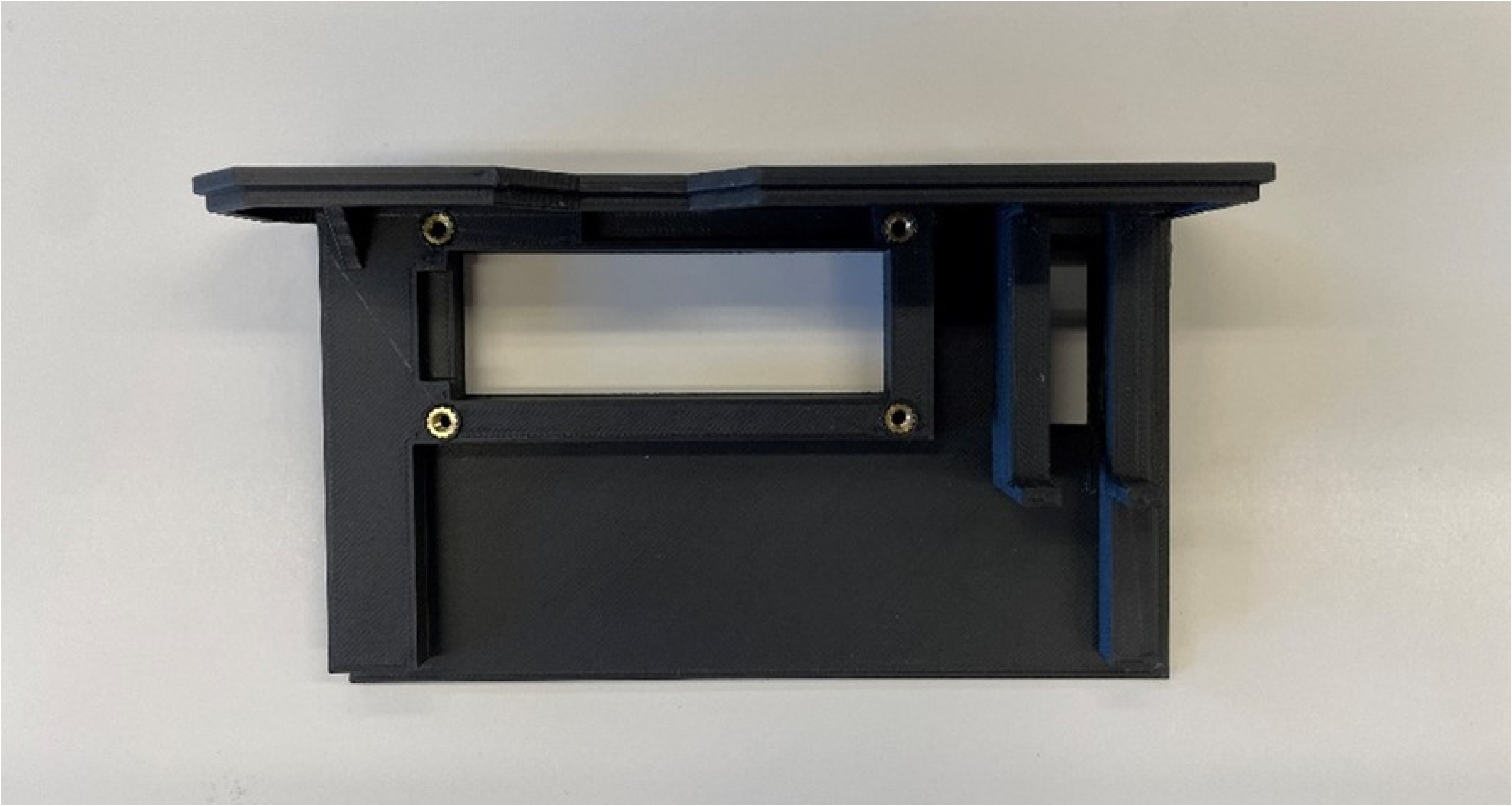
I2C Adapter to LCD screen pin connect

**Figure 24.**
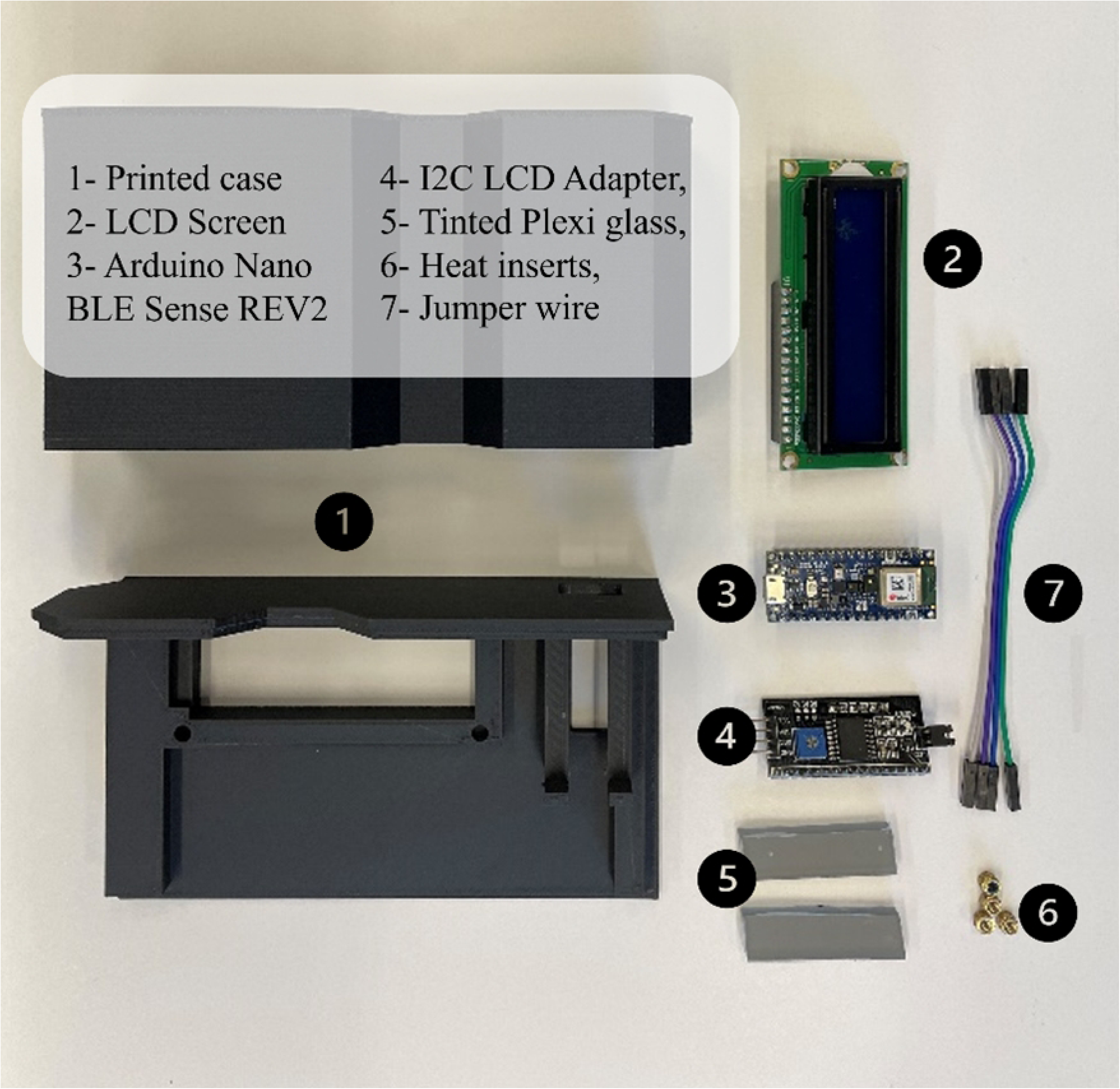
Screen in place with wires.

**Figure 25.**
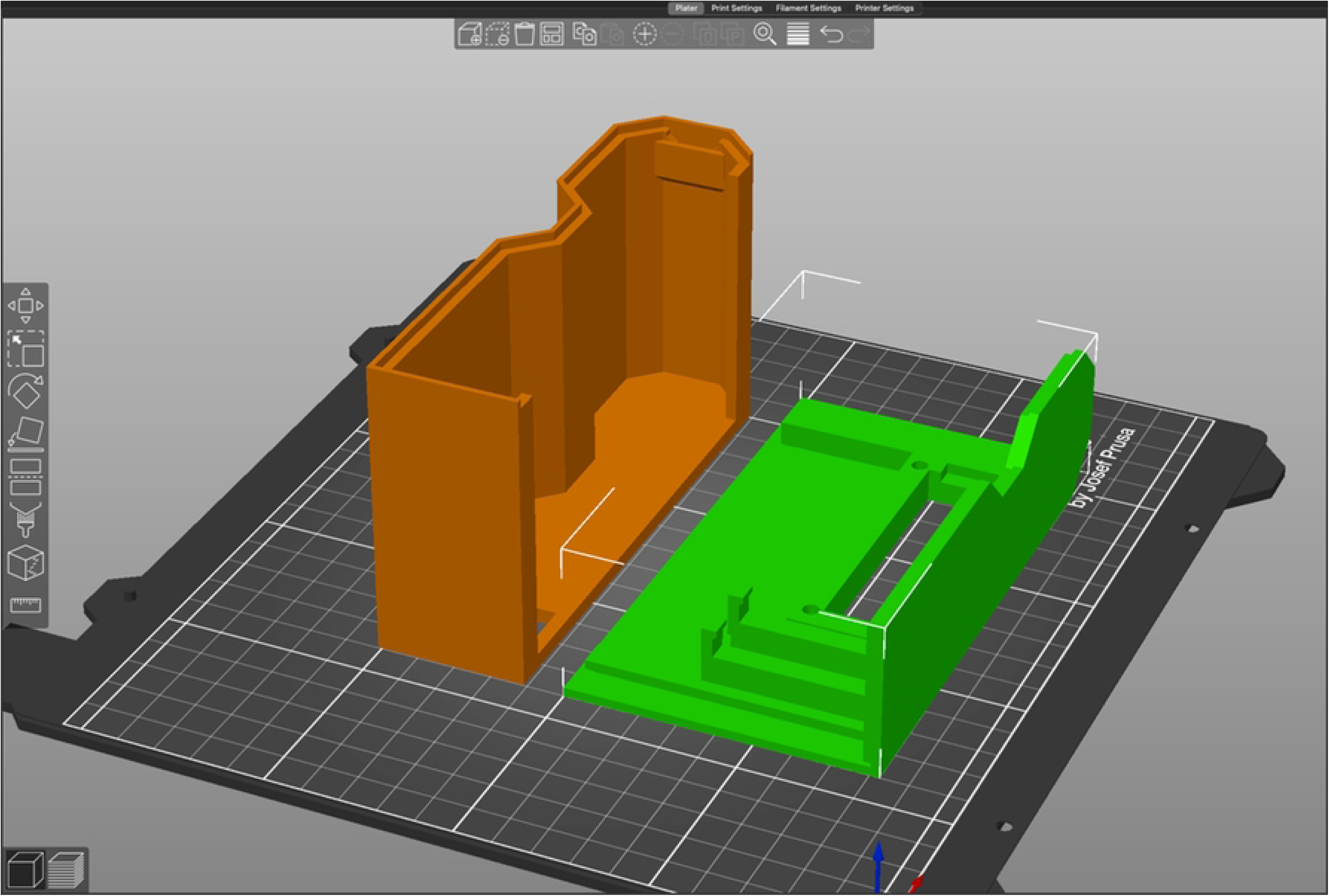
AS7256x Sensor being mounted.

**Figure 26.**
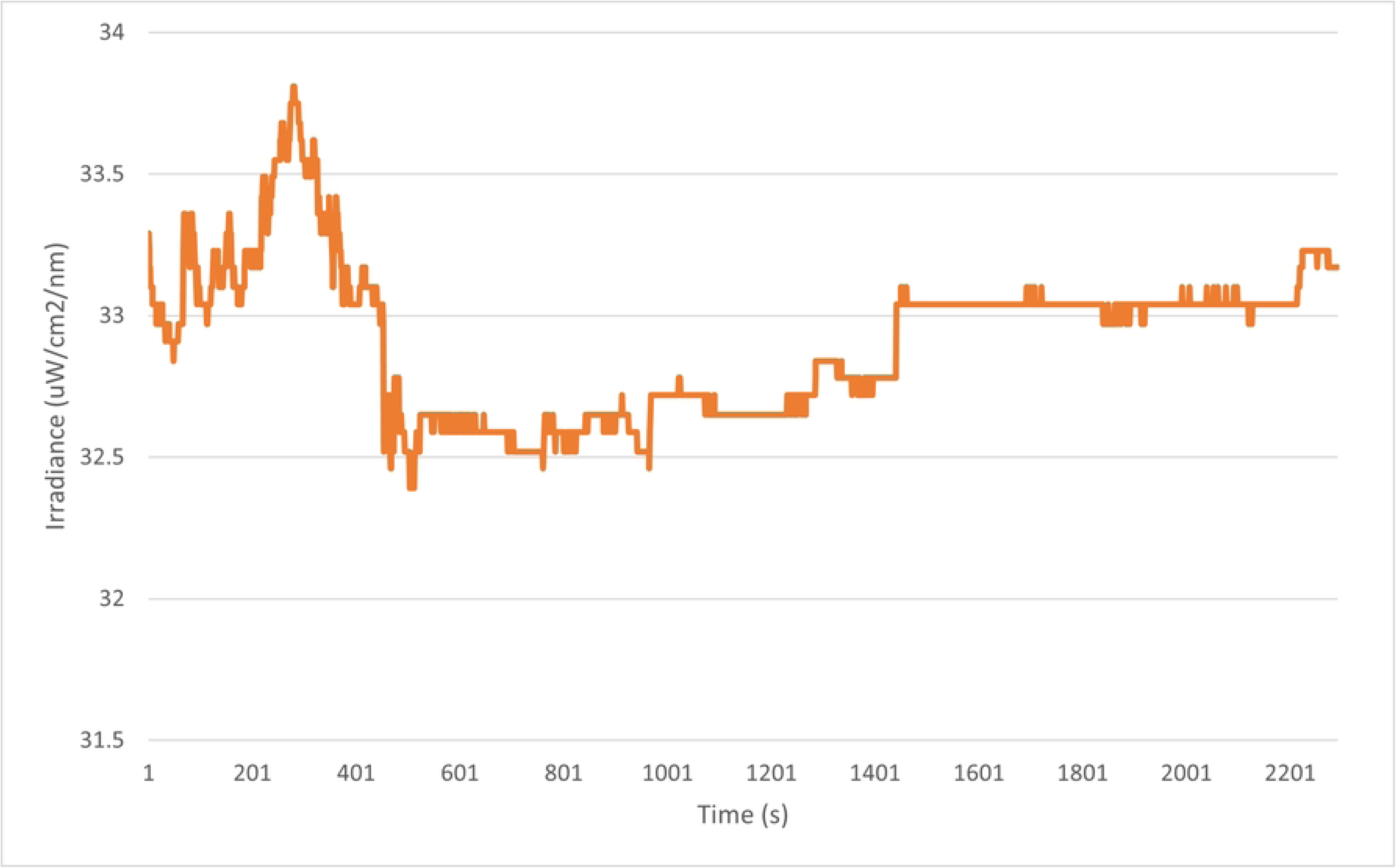
Single layer of scotch tape placed in either side.

**Figure 27.**
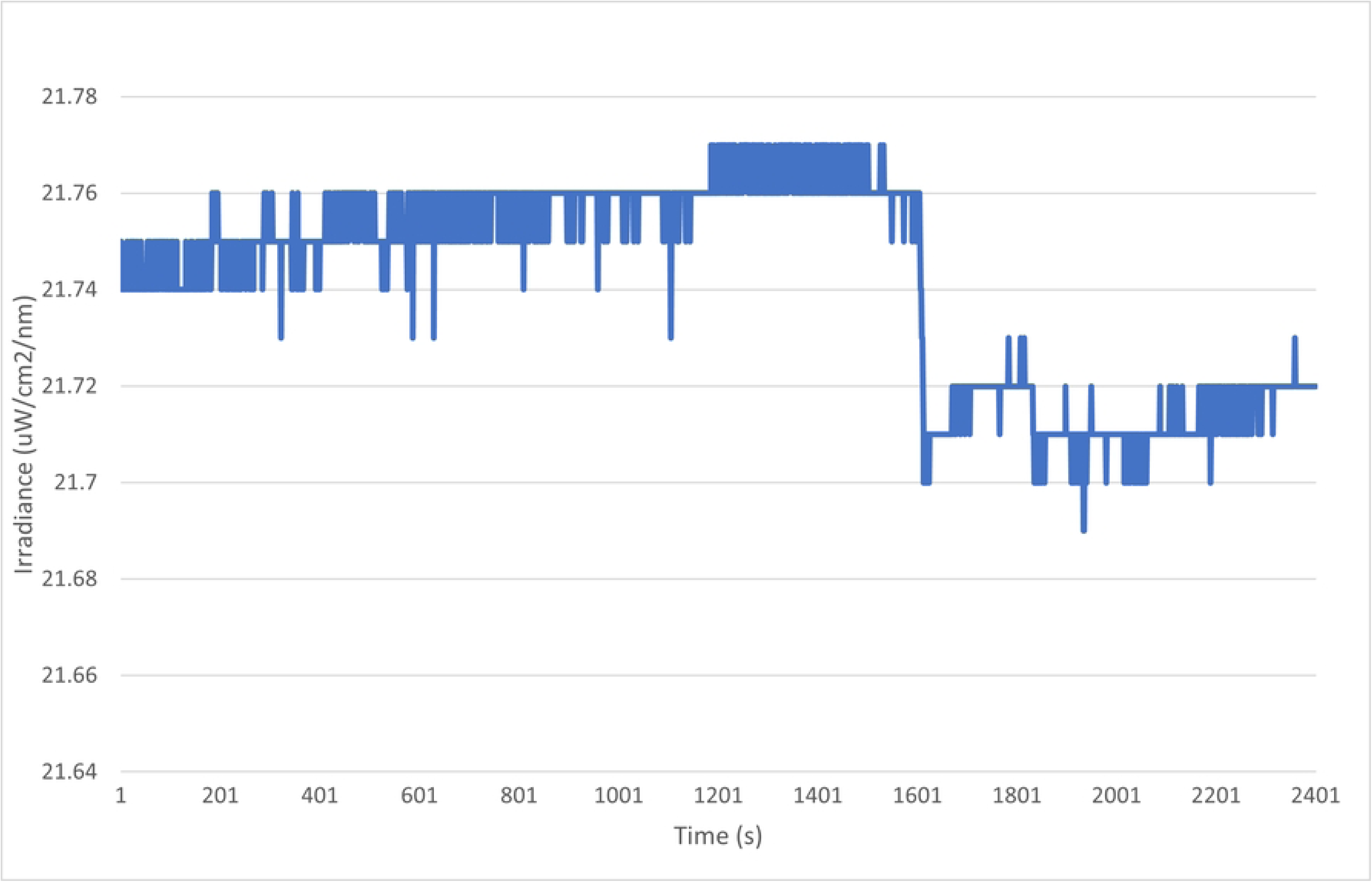
Comparison of plexiglass with tape and without

**Figure 28.**
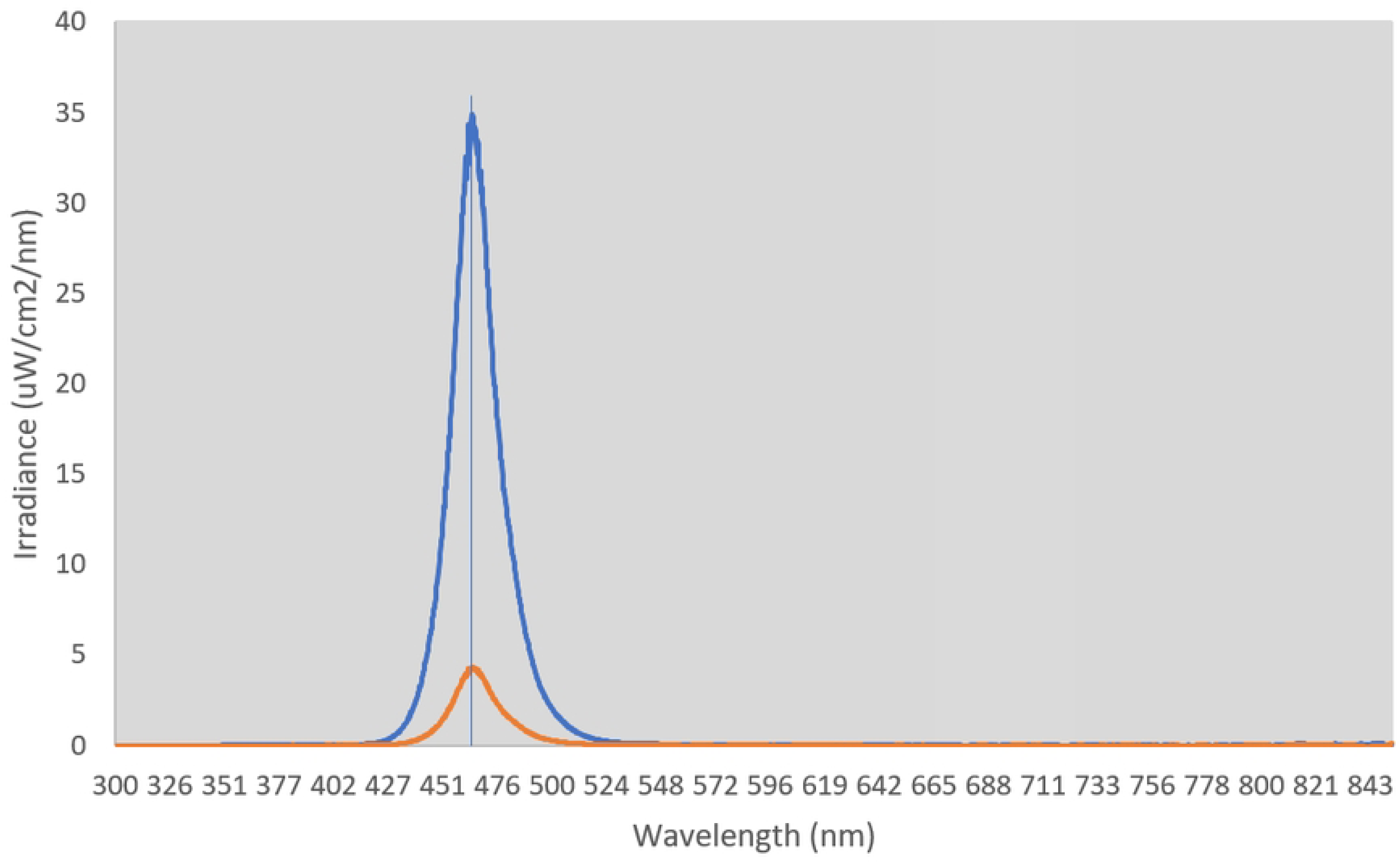
Filter in place

**Figure 29.**
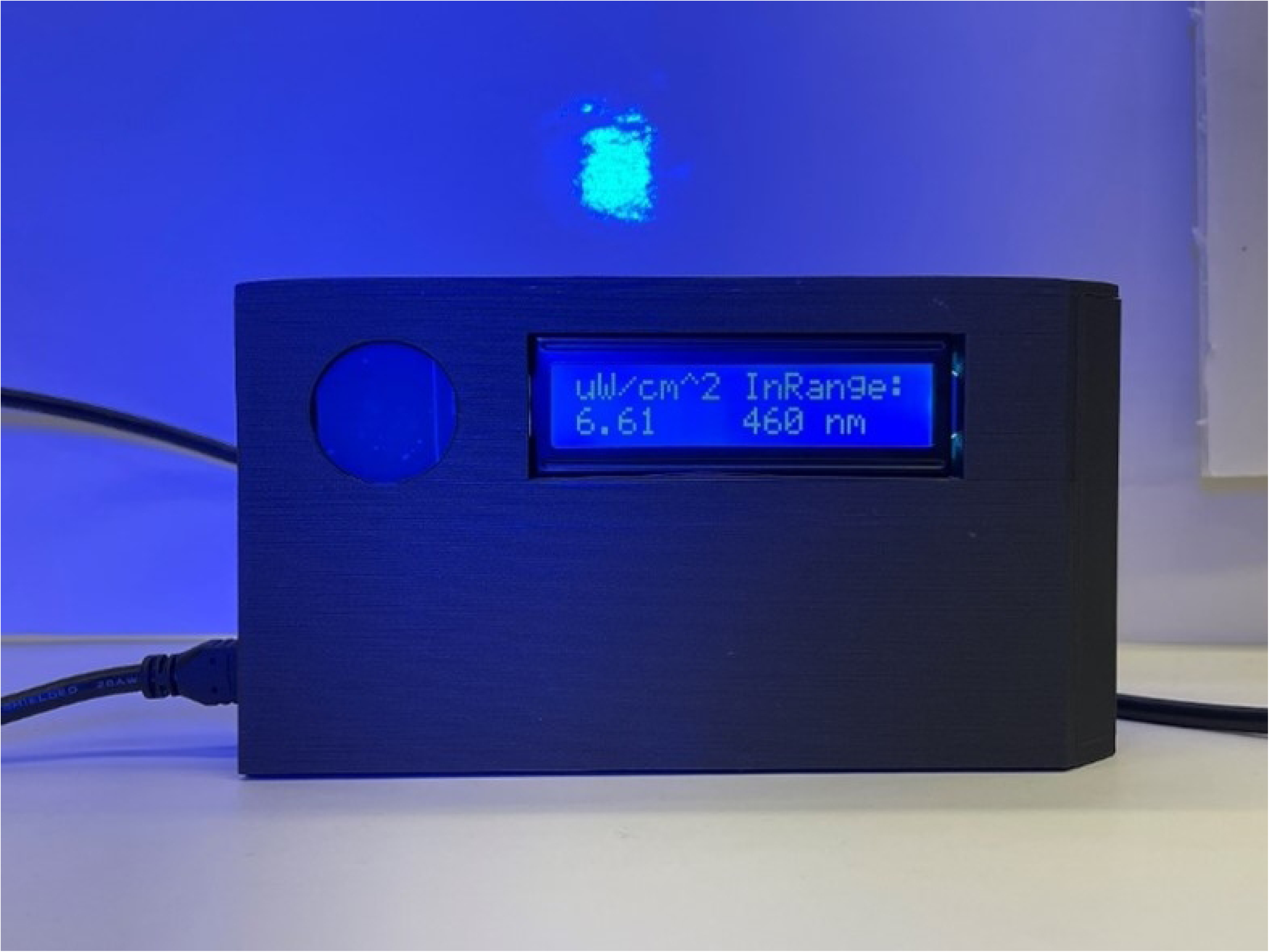
5V pin activated.

**Figure 30.**
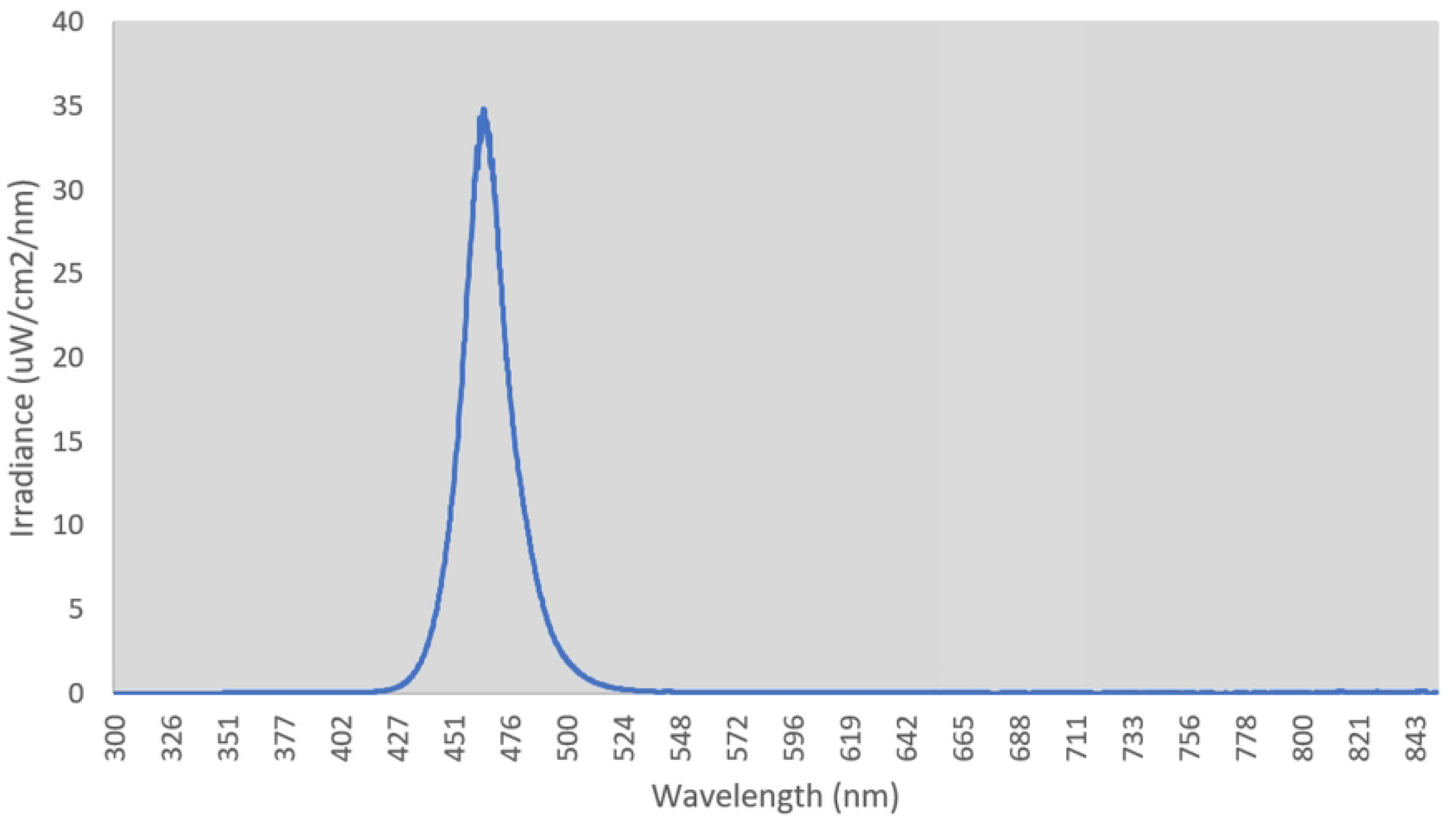
Top side of board and pins

**Figure 31.**
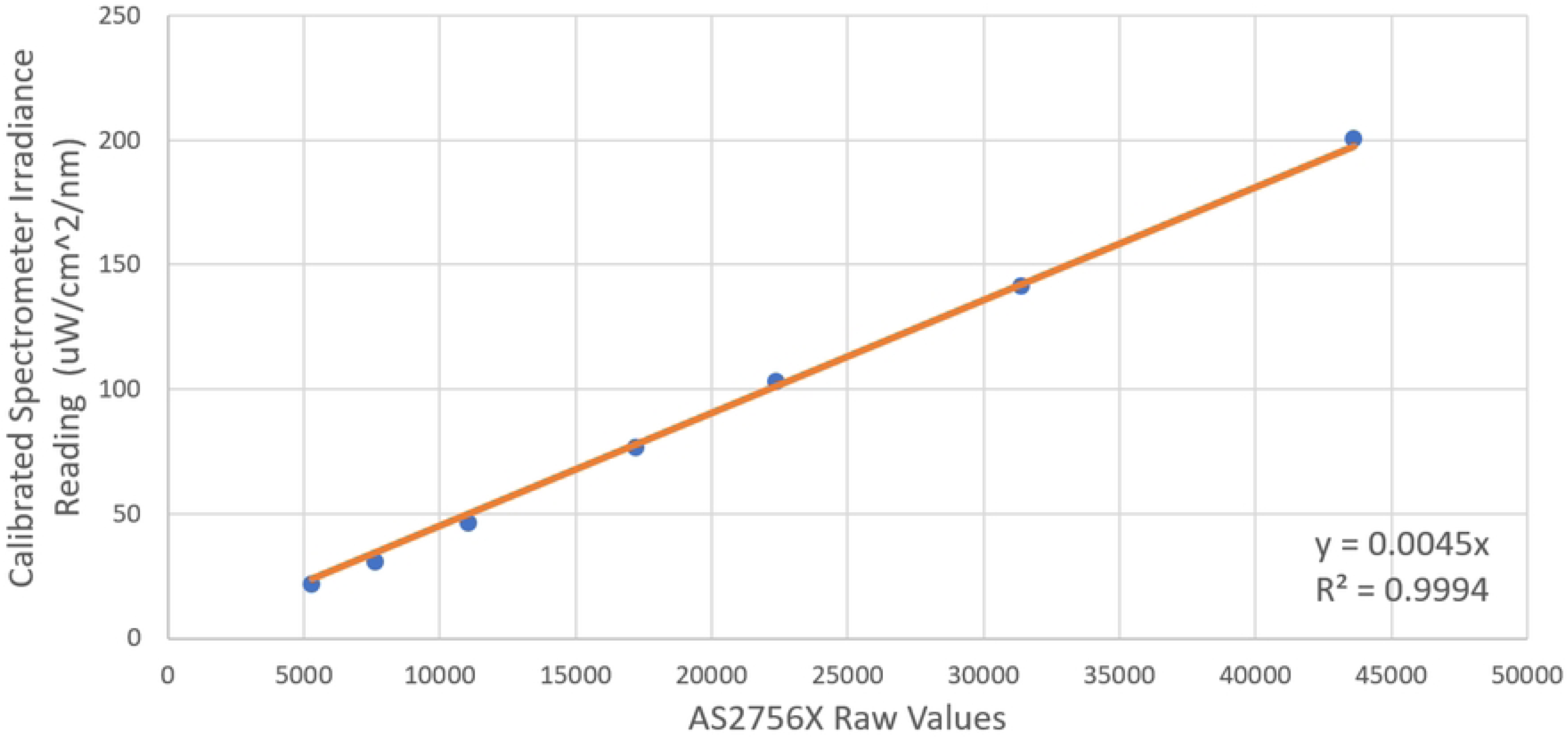
Reverse side of board and pins

**Figure 32.**
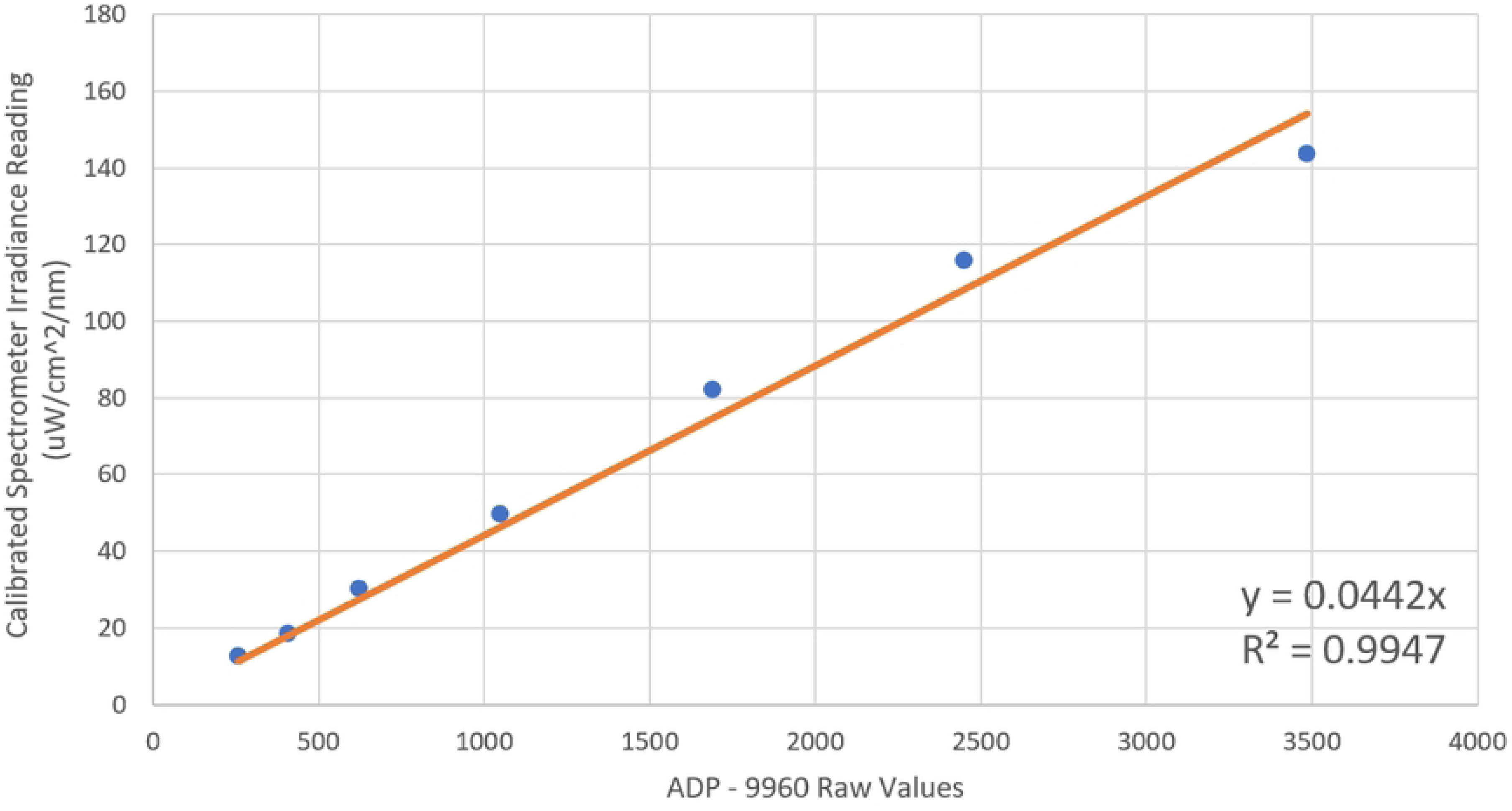
Completed device internals.

**Figure 33.**
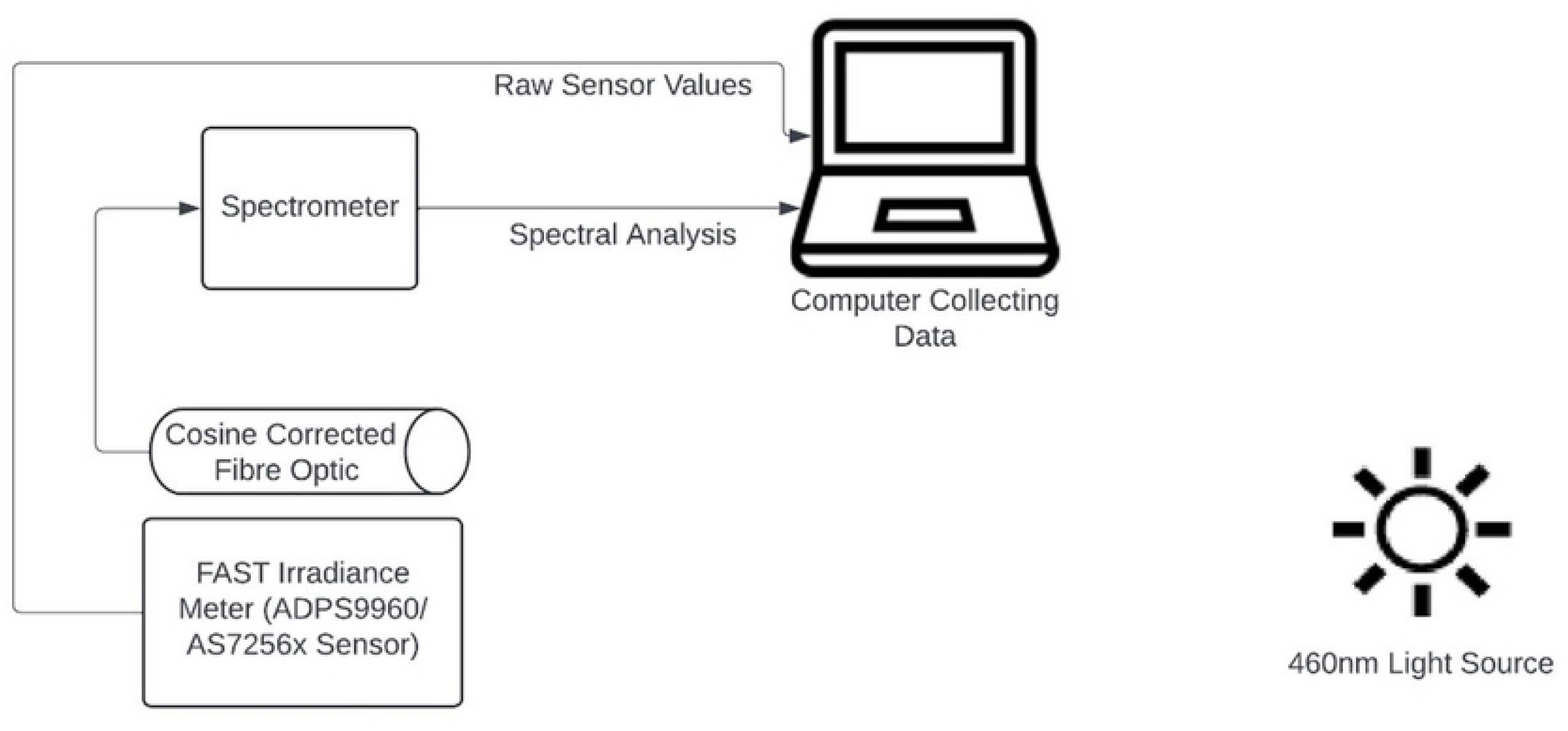
Irradiance Meter displaying calibrated results

**Figure 34.**
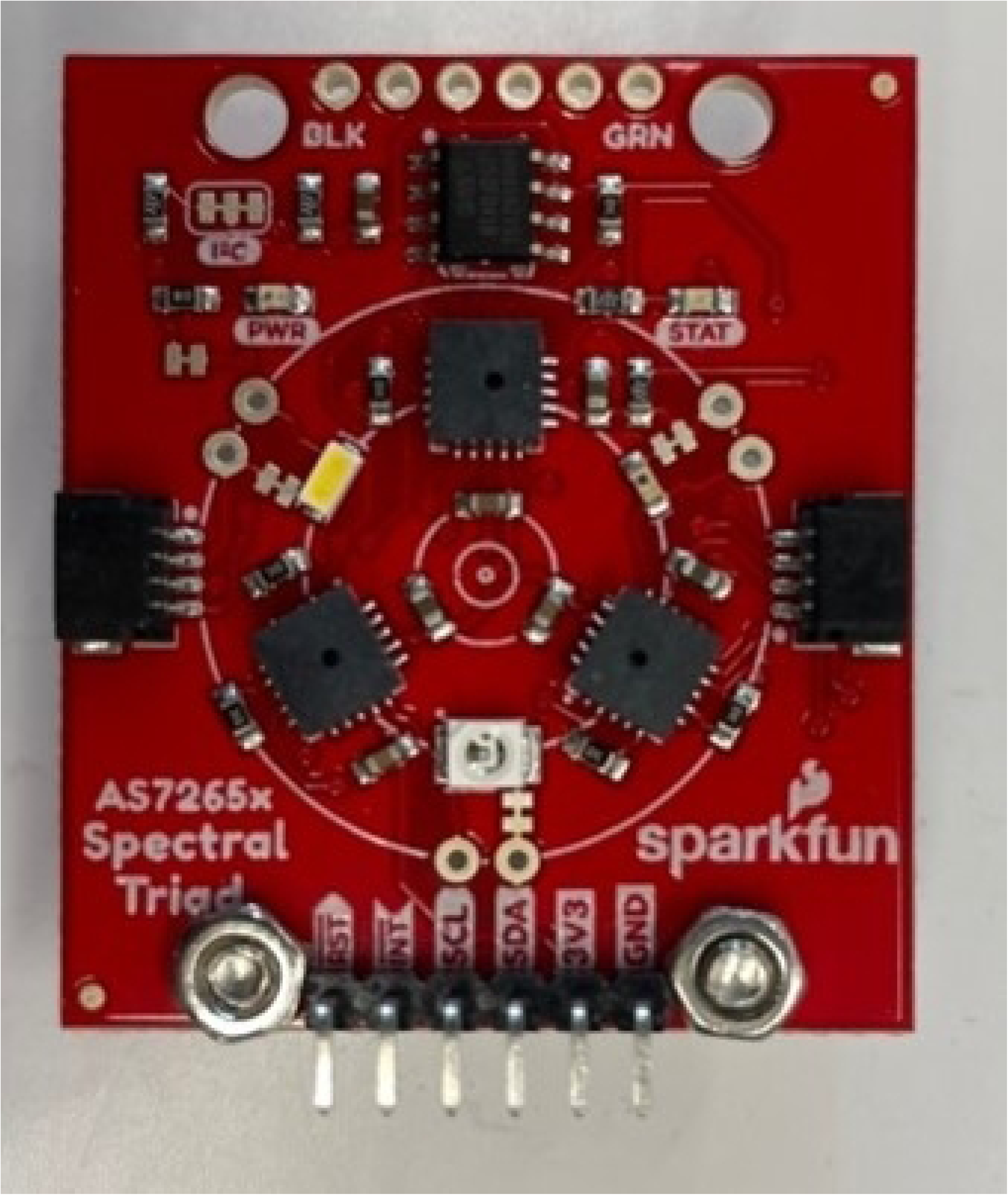
Unassembled Battery Pack Variation

**Figure 35.**
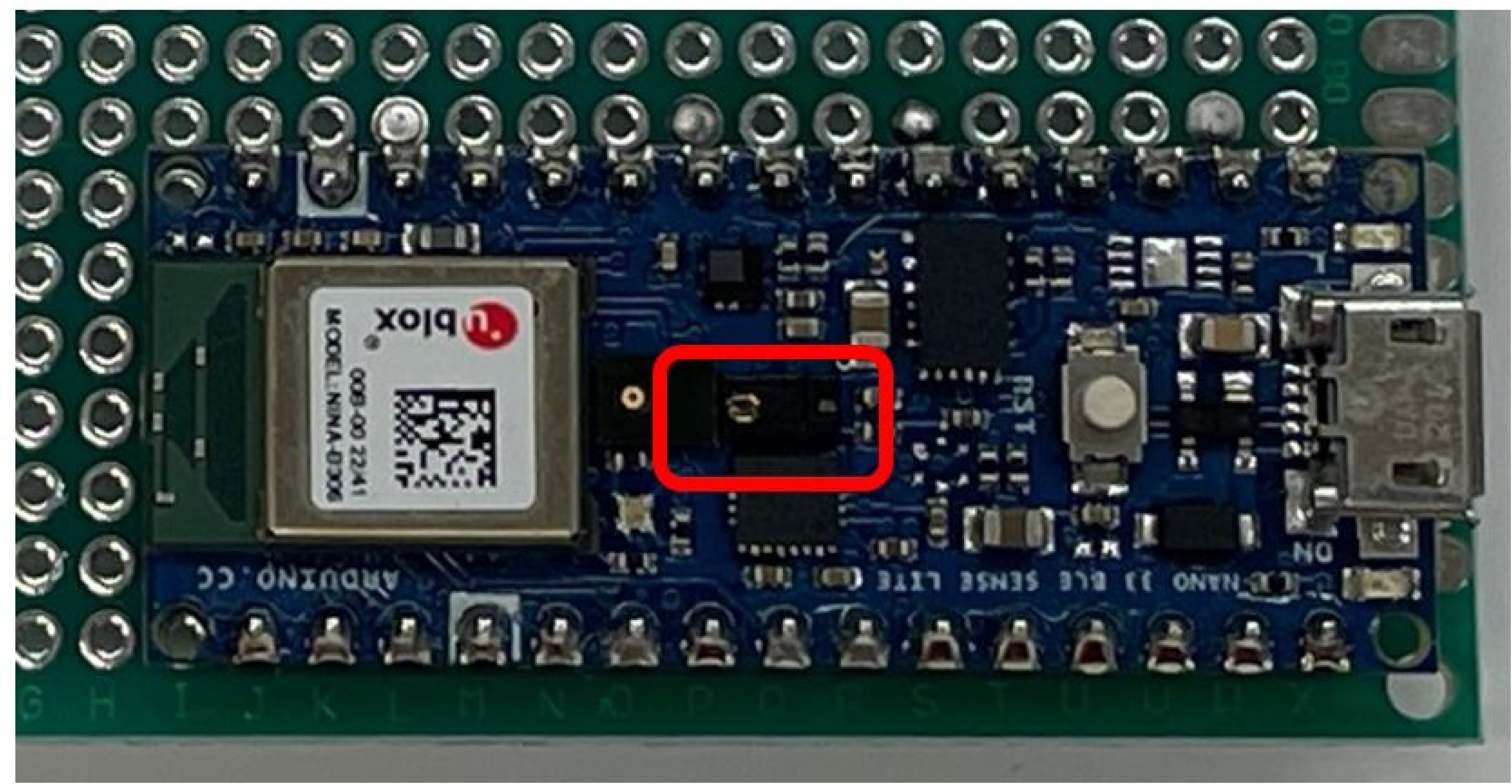
Assembled Battery Operated AS2756X Irradiance meter

## References

1. Porter ML, Dennis BL. Hyperbilirubinemia in the Term Newborn. Am Fam Physician. 2002 Feb 15;65(4):599–607.

2. Olusanya BO, Osibanjo FB, Slusher TM. Risk Factors for Severe Neonatal Hyperbilirubinemia in Low and Middle-Income Countries: A Systematic Review and Meta-Analysis. Carlo WA, editor. PLOS ONE. 2015 Feb 12;10(2):e0117229.

3. Ansong-Assoku B, Shah SD, Adnan M, Ankola PA. Neonatal Jaundice. In: StatPearls [Internet]. Treasure Island (FL): StatPearls Publishing; 2023 [cited 2023 Aug 14]. Available from: http://www.ncbi.nlm.nih.gov/books/NBK532930/

4. Slusher TM, Zamora TG, Appiah D, Stanke JU, Strand MA, Lee BW, et al. Burden of severe neonatal jaundice: a systematic review and meta-analysis. BMJ Paediatr Open. 2017 Nov;1(1):e000105.

5. Bhutani VK, Zipursky A, Blencowe H, Khanna R, Sgro M, Ebbesen F, et al. Neonatal hyperbilirubinemia and Rhesus disease of the newborn: incidence and impairment estimates for 2010 at regional and global levels. Pediatr Res. 2013 Dec;74(S1):86–100.

6. Muchowski KE. Evaluation and Treatment of Neonatal Hyperbilirubinemia. Am Fam Physician. 2014 Jun 1;89(11):873–8.

7. Mohan DR, Lu H, McClary J, Marasch J, Nock ML, Ryan RM. Evaluation of Intravenous Immunoglobulin Administration for Hyperbilirubinemia in Newborn Infants with Hemolytic Disease. Children. 2023 Mar 2;10(3):496.

8. Zhang M, Tang J, He Y, Li W, Chen Z, Xiong T, et al. Systematic review of global clinical practice guidelines for neonatal hyperbilirubinemia. BMJ Open. 2021 Jan;11(1):e040182.

9. Bhutani VK, the Committee on Fetus and Newborn. Phototherapy to Prevent Severe Neonatal Hyperbilirubinemia in the Newborn Infant 35 or More Weeks of Gestation. Pediatrics. 2011 Oct 1;128(4):e1046–52.

10. Cline BK, Vreman HJ, Faber K, Lou H, Donaldson KM, Amuabunosi E, et al. Phototherapy Device Effectiveness in Nigeria: Irradiance Assessment and Potential for Improvement. J Trop Pediatr. 2013 Aug 1;59(4):321–5.

11. Tan KL. Comparison of the effectiveness of phototherapy and exchange transfusion in the management of nonhemolytic neonatal hyperbilirubinemia. J Pediatr. 1975 Oct;87(4):609–12.

12. Abe S, Fujioka K. Can exchange transfusion be replaced by double-LED phototherapy? Open Med. 2021 Jul 2;16(1):992–6.

13. Kumar P, Chawla D, Deorari A. Light-emitting diode phototherapy for unconjugated hyperbilirubinaemia in neonates. Cochrane Neonatal Group, editor. Cochrane Database Syst Rev [Internet]. 2011 Dec 7 [cited 2023 Aug 21]; Available from: https://doi.wiley.com/10.1002/14651858.CD007969.pub2

14. Wentworth SD. Neonatal phototherapy–today’s lights, lamps and devices. Infant. 2005;1(1):14– 9.

15. Wang J, Guo G, Li A, Cai WQ, Wang X. Challenges of phototherapy for neonatal hyperbilirubinemia (Review). Exp Ther Med. 2021 Jan 20;21(3):231.

16. Sisson TRC. Visible Light Therapy of Neonatal Hyperbilirubinemia. In: Smith KC, editor. Photochemical and Photobiological Reviews: Volume 1 [Internet]. Boston, MA: Springer US; 1976 [cited 2023 Aug 1]. p. 241–68. Available from: 10.1007/978-1-4684-2574-1_6

17. Sisson TRC. Molecular Basis of Hyperbilirubinemia and Phototherapy. J Invest Dermatol. 1981 Jul;77(1):158–61.

18. Stokowski LA. Fundamentals of Phototherapy for Neonatal Jaundice. Adv Neonatal Care. 2011 Oct;11(Supplement 5S):S10–21.

19. phototherapy-light-TPP.pdf [Internet]. [cited 2023 Aug 19]. Available from: https://www.unicef.org/supply/media/2926/file/phototherapy-light-TPP.pdf

20. Sampurna MTA, Etika R, Utomo MT, Rani SAD, Irzaldy A, Irawan ZS, et al. An evaluation of phototherapy device performance in a tertiary health facility. Heliyon. 2020 Sep;6(9):e04950.

21. Phototherapy irradiance meter [Internet]. [cited 2024 Feb 18]. Available from: https://supply.unicef.org/s0002018.html

22. Phototherapy & Irradiance Radiometers | Fluke Biomedical [Internet]. [cited 2024 Feb 18]. Available from: https://www.flukebiomedical.com/products/biomedical-test-equipment/phototherapy-radiometers

23. ILT750 Bili Light Meter/Radiometer for Neonatal Jaundice Treatment [Internet]. [cited 2024 Feb 18]. Available from: https://internationallight.com/products/ilt750-bili-light-meter

24. Mayowa A, Abioye Abiodun E. Design and analysis of a solar powered phototherapy device. J Phys Conf Ser. 2019 Dec 1;1378(3):032041.

25. Pearce JM. Building Research Equipment with Free, Open-Source Hardware. Science [Internet]. 2012 Sep 14 [cited 2023 Sep 23];337(6100):1303–4. Available from: https://www.science.org/doi/abs/10.1126/science.1228183

26. Pearce JM. Open-Source Lab: How to Build Your Own Hardware and Reduce Research Costs. Elsevier; 2013. 291 p.

27. Pearce JM. Cut costs with open-source hardware. Nature [Internet]. 2014 Jan [cited 2023 Sep 18];505(7485):618–618. Available from: https://www.nature.com/articles/505618d

28. Coakley MF, Hurt DE, Weber N, Mtingwa M, Fincher EC, Alekseyev V, et al. The NIH 3D Print Exchange: A Public Resource for Bioscientific and Biomedical 3D Prints. 3D Print Addit Manuf [Internet]. 2014 Sep [cited 2023 Oct 23];1(3):137–40. Available from: https://www.liebertpub.com/doi/abs/10.1089/3Dp.2014.1503

29. Pearce JM. Distributed Manufacturing of Open Source Medical Hardware for Pandemics. J Manuf Mater Process [Internet]. 2020 Jun [cited 2023 Feb 9];4(2):49. Available from: https://www.mdpi.com/2504-4494/4/2/49

30. De Maria C, Di Pietro L, Ravizza A, Lantada AD, Ahluwalia AD. Chapter 2 Open-source medical devices: Healthcare solutions for low-, middle-, and high-resource settings. In: Iadanza E, editor. Clinical Engineering Handbook (Second Edition) [Internet]. Academic Press; 2020 [cited 2023 Jul 25]. p. 7–14. Available from: https://www.sciencedirect.com/science/article/pii/B978012813467200002X

31. Gibb A. Building Open Source Hardware: DIY Manufacturing for Hackers and Makers. Addison-Wesley Professional; 2014. 369 p.

32. Oberloier S, Pearce JM. General Design Procedure for Free and Open-Source Hardware for Scientific Equipment. Designs [Internet]. 2018 Mar [cited 2023 Jun 14];2(1):2. Available from: https://www.mdpi.com/2411-9660/2/1/2

33. Pearce JM. Economic savings for scientific free and open source technology: A review. HardwareX [Internet]. 2020 Oct 1 [cited 2023 Jun 14];8:e00139. Available from: https://www.sciencedirect.com/science/article/pii/S2468067220300481

34. Maia Chagas A. Haves and have nots must find a better way: The case for open scientific hardware. PLOS Biol [Internet]. 2018 Sep 27 [cited 2023 Jun 15];16(9):e3000014. Available from: https://dx.plos.org/10.1371/journal.pbio.3000014

35. Otero J, Pearce JM, Gozal D, Farré R. Open-source design of medical devices. Nat Rev Bioeng [Internet]. 2024 Feb 15 [cited 2024 Mar 16];1–2. Available from: https://www.nature.com/articles/s44222-024-00162-9

36. SR2 UV-VIS Spectrometers [Internet]. [cited 2024 Feb 18]. Available from: https://www.oceaninsight.com/products/spectrometers/general-purpose-spectrometer/ocean-sr2-series-spectrometers/ocean-sr2-uv-vis-spectrometers/

37. Datex-Ohmeda Giraffe Spot PT Lite Phototherapy System Buy, Rent, or Lease [Internet]. [cited 2024 May 12]. Available from: https://www.medonegroup.com/equipment/therapy/datex-ohmeda-giraffe-spot-pt-lite-phototherapy-system

38. Avago-APDS-9960-datasheet.pdf [Internet]. [cited 2024 Apr 6]. Available from: https://cdn.sparkfun.com/assets/learn_tutorials/3/2/1/Avago-APDS-9960-datasheet.pdf

39. Arduino Online Shop [Internet]. [cited 2024 Feb 28]. Arduino Nano 33 BLE Sense. Available from: https://store-usa.arduino.cc/products/arduino-nano-33-ble-sense

40. SparkFun Triad Spectroscopy Sensor AS7265x (Qwiic) SEN-15050 SparkFun Electronics [Internet]. [cited 2024 Feb 18]. Available from: https://www.sparkfun.com/products/15050

41. Ocean View [Internet]. Ocean Insight; Available from: https://www.oceaninsight.com/products/software/acquisition-and-analysis/oceanview/

42. Ocean Insight Spectroscopy Software for Download [Internet]. [cited 2024 Feb 18]. Available from: https://www.oceaninsight.com/support/software-downloads/

43. Snapklik.com [Internet]. [cited 2024 Apr 4]. Snapklik.com : BuyPlastic Polycarbonate Plastic Sheet 7130 Gray. Available from: https://snapklik.com/en-ca/product/09WH4PM7P1OU5

44. Scotch® Magic^TM^ Invisible Tape, 810D, with refillable dispenser, 3/4 in x 36 yd (19 mm x 33 m) [Internet]. [cited 2024 Apr 4]. Available from: https://www.3mcanada.ca/3M/en_CA/p/d/v000074913/

45. Lightmeter / 4meter – LifeKit by MTTS [Internet]. [cited 2025 Jul 1]. Available from: https://www.mtts-asia.com/lightmeter/

46. Draeger. Photo-Therapy 4000 Jaundice Management.

47. Phototherapy irradiance meter [Internet]. [cited 2024 Feb 18]. Available from: https://supply.unicef.org/s0002018.html

48. Zaaba NF, Jaafar M. A review on degradation mechanisms of polylactic acid: Hydrolytic, photodegradative, microbial, and enzymatic degradation. Polym Eng Sci. 2020 Sep;60(9):2061–75.

49. Jones R, Haufe P, Sells E, Iravani P, Olliver V, Palmer C, et al. RepRap – the replicating rapid prototyper. Robotica [Internet]. 2011 Jan [cited 2023 Aug 11];29(1):177–91. Available from: https://www.cambridge.org/core/journals/robotica/article/reprap-the-replicating-rapid-prototyper/5979FD7B0C066CBCE43EEAD869E871AA

50. Sells E, Bailard S, Smith Z, Bowyer A, Olliver V. RepRap: The Replicating Rapid Prototyper: Maximizing Customizability by Breeding the Means of Production. In: Handbook of Research in Mass Customization and Personalization [Internet]. World Scientific Publishing Company; 2009 [cited 2023 Aug 11]. p. 568–80. Available from: https://www.worldscientific.com/doi/abs/10.1142/9789814280280_0028

51. Bowyer A. 3D Printing and Humanity’s First Imperfect Replicator. 3D Print Addit Manuf [Internet]. 2014 Mar [cited 2023 Aug 11];1(1):4–5. Available from: https://www.liebertpub.com/doi/abs/10.1089/3dp.2013.0003

52. Givans J. FAST_IrradianceMeter. 2024 [cited 2024 May 3]; Available from: https://osf.io/7dqp6/

